# Diagnostic accuracy of three prevailing rapid antigen tests for detection of SARS-CoV-2 infection in the general population: cross sectional study

**DOI:** 10.1101/2021.11.19.21266579

**Authors:** Roderick P Venekamp, Irene K Veldhuijzen, Karel G M Moons, Wouter van den Bijllaardt, Suzan D Pas, Esther B Lodder, Richard Molenkamp, Zsofi Igloi, Constantijn Wijers, Claudy Oliveira dos Santos, Sylvia B Debast, Marjan J. Bruins, Khaled Polad, Carla R S Nagel-Imming, Wanda G H Han, Janneke H H M van de Wijgert, Susan van den Hof, Ewoud Schuit

**Affiliations:** Julius Center for Health Sciences and Primary Care, University Medical Center Utrecht, Utrecht University, Utrecht, The Netherlands; Centre for Infectious Disease Control, National Institute for Public Health and the Environment (RIVM), Bilthoven, The Netherlands; Cochrane Netherlands, University Medical Center Utrecht, Utrecht University, Utrecht, The Netherlands; Microvida Laboratory for Medical Microbiology, Amphia Hospital, Breda, The Netherlands; Microvida Laboratory for Medical Microbiology, Bravis Hospital, Roosendaal, The Netherlands; Public Health Service West-Brabant, Breda, The Netherlands; Department of Viroscience, Erasmus MC, Rotterdam, The Netherlands; Public Health Service Rotterdam-Rijnmond, Rotterdam, The Netherlands; Laboratory of Medical Microbiology and Infectious Diseases, Isala Hospital, Zwolle, The Netherlands; Public Health Service IJsselland, Zwolle, The Netherlands

## Abstract

**Objective:** To assess the diagnostic accuracy of three rapid antigen tests (Ag-RDTs) for detecting SARS-CoV-2 infection in the general population.

**Design:** Cross-sectional study with follow-up using pseudonymised record linkage.

**Setting:** Three Dutch public health service COVID-19 test sites.

**Participants:** Consecutively included individuals aged 16 years and older presenting for SARS-CoV-2 testing.

**Main outcome measures:** Sensitivity, specificity, positive and negative predictive values of *BD-Veritor*^*tm*^ *System (Becton Dickinson*), *PanBio (Abbott*), and *SD-Biosensor (Roche Diagnostics*), applying routinely used sampling methods (combined oropharyngeal and nasal [OP-N] or nasopharyngeal [NP] swab), with molecular testing as reference standard. For SD-Biosensor, the diagnostic accuracy with OP-N sampling was also assessed. A viral load cut-off (≥5.2 log10 SARS-CoV-2 E-gene copies/mL) served as a proxy of infectiousness.

**Results:** SARS-CoV-2 prevalence and overall sensitivities with 95% confidence intervals were 188/1441 (13.0%) and 129/188 (68.6% [61.5%-75.2%]) for BD-Veritor, 173/2056 (8.4%) and 119/173 (68.8% [61.3%-75.6%]) for PanBio, and 215/1769 (12.2%) and 160/215 (74.4% [68.0%-80.1%]) for SD-Biosensor with routine sampling, and 164/1689 (9.7%) and 123/164 (75.0% [67.7%-81.4%]) for SD-Biosensor with OP-N sampling. In those symptomatic or asymptomatic at sampling, sensitivities were 72.2%-83.4% and 54.0%-55.9%, respectively. With a viral load cut-off, sensitivities were 125/146 (85.6% [78.9%-90.9%]) for BD-Veritor, 108/121 (89.3% [82.3%-94.2%]) for PanBio, 160/182 (87.9% [82.3%-92.3%]) for SD-Biosensor with routine sampling, and 118/141 (83.7% [76.5%-89.4%]) with OP-N sampling. Specificities were >99%, and positive and negative predictive values >95%, for all tests in most analyses. 61.3% of false negative Ag-RDT participants returned for testing within 14 days (median of 3 days, interquartile range 3) of whom 90.3% tested positive.

**Conclusions:** The overall sensitivities of the three Ag-RDTs were 68.6%-75.0%, increasing to at least 85.6% after the viral load cut-off was applied. For SD-Biosensor, the diagnostic accuracy with OP-N and NP sampling was comparable. Over 55% of false negative Ag-RDT participants tested positive during follow-up.

## Introduction

In the first phase of the pandemic, all testing at Dutch public health service COVID-19 test sites was done with molecular tests. A molecular test, mainly real-time reverse-transcriptase polymerase chain reaction (RT-PCR), is still considered the reference test for SARS-CoV-2.^1^ However, molecular testing platforms are typically only available in centralised laboratories and most of them require sample batching, thereby causing delays in delivering test results. Persons with symptoms were – and still are – strongly advised to isolate themselves until a negative test result has been obtained. Reducing test-to-result delays is therefore considered important. Point-of-care antigen tests have that potential and were introduced for testing of symptomatic persons at Dutch public health service test sites in November 2020. Later, these were also introduced for testing of close contacts who were asymptomatic at the time of sampling, to gain entry to places and events where physical distancing is difficult to achieve or enforce, for travel, and for self-testing at home. Rapid lateral flow antigen diagnostic tests (Ag-RDTs) are the most promising and widely used point-of-care tests.^2^ They require no or minimal equipment, provide a result within 15 minutes, and can be performed in a range of settings. Even in countries with high COVID-19 vaccination coverage, Ag-RDTs are expected to continue to play a pivotal role as countries are reopening and physical distancing measures are relaxed.

Thus far, multiple studies investigated the diagnostic accuracy of SARS-CoV-2 point-of-care tests.^3-5^ However, most of these studies had a limited sample size, used specimens that were left-over after molecular testing, or included symptomatic individuals only. We conducted a large diagnostic accuracy study in late 2020/early 2021 in which two SARS-CoV-2 Ag-RDTs (*BD Veritor*^*tm*^ *System by Becton Dickinson* (‘BD-Veritor’) and *Roche/SD Biosensor by Roche Diagnostics* (‘SD-Biosensor’)) were compared to RT-PCR.^6^ However, we limited that evaluation to asymptomatic and presymptomatic close contacts of individuals with confirmed SARS-CoV-2 infection and one of the most commonly used Ag-RDTs in The Netherlands, *PanBio by Abbott* (‘PanBio’), was not included in that study. Furthermore, the diagnostic accuracy of Ag-RDTs may alter over time due to changes in SARS-CoV-2 epidemiology and indications for testing, and roll-out of COVID-19 vaccination. Diagnostic accuracy may also be impacted by sampling technique. Many Ag-RDTs require deep nasopharyngeal (NP) sampling, which is often considered to be unpleasant, whereas oropharyngeal combined with superficial nasal (OP-N) sampling might suffice.

In the first phase of the current study, we therefore evaluated the diagnostic accuracies of three Ag-RDTs (BD-Veritor, SD-Biosensor, and PanBio) that are commonly applied in Dutch test sites, using the sampling techniques that are routinely used by those test sites in individuals aged 16 or older irrespective of their indication for testing, symptomatology, and COVID-19 vaccination status. In the second phase of the study, we also evaluated the diagnostic accuracies of two of the Ag-RDTs (SD-Biosensor and PanBio) when using a less invasive OP-N sampling technique.

## Methods

The study is reported according to the STARD 2015 guidelines: an updated list of essential items for reporting diagnostic accuracy studies.^7^

### Study design and population

This large cross-sectional diagnostic test accuracy study was embedded within the Dutch public testing infrastructure. Public testing in the Netherlands is free-of-charge but only available for government-approved test indications. At the time of the study (12 April to 14 June 2021), these indications included having symptoms of suspected SARS-CoV-2 infection or having been identified as a close contact of a SARS-CoV-2 index case via traditional contact-tracing or the contact-tracing app regardless of symptomatology at the time of notification. Participants were recruited consecutively at three Dutch public health service COVID-19 test sites across the country, located in the West-Brabant region (Breda, using the BD-Veritor Ag-RDT), in the Rotterdam-Rijnmond region (Rotterdam The Hague Airport and Ahoy, using the SD-Biosensor Ag-RDT) and the IJsselland region (Zwolle, using the PanBio Ag-RDT). Individuals were considered eligible if they were aged 16 years or older, and willing and able to sign an informed consent in Dutch.

The Dutch COVID-19 vaccination programme started on 6 January 2021. At the time of the study, an estimated 20% (12 April) to 62% (14 June) of Dutch inhabitants aged 18 or older had received at least one vaccination, ranging over time from 4%-13% for 18-25 year olds to 85%-91% for 81-90 year olds.^8^ The study was conducted before the SARS-CoV-2 Delta variant became the dominant variant in the Netherlands in June 2021 (the prevalence of the Delta variant was around 8.6% in the last week of inclusions).^9,10^

### Inclusion procedure

Participants arrived at the test sites by car or bicycle (Breda) or on foot (Rotterdam and Zwolle). Test site staff verbally verified study eligibility. Eligible individuals were given a study flyer and a participant information letter to read, after which they could indicate to site staff if they wanted to participate. After signing the informed consent form, participants completed a short questionnaire on indication for testing, presence, type, and onset of symptoms, previous SARS-CoV-2 infections, and COVID-19 vaccination status (supplementary material 1) while waiting for sampling.

### Specimen collection and testing

A trained test site staff member took two swabs from each study participant: one for molecular reference testing and the other for the Ag-RDT. The molecular reference test was performed in a centralised laboratory in each region, whereas the Ag-RDT was performed at the test sites. In the first phase of the study, the Ag-RDT swabs were collected using the sampling method that was routinely used at the test site, i.e., deep NP for SD-Biosensor and PanBio, and superficial OP-N (about 2.5 cm deep) for BD-Veritor. In the second phase of the study, we evaluated the SD-Biosensor and PanBio tests using less invasive OP-N sampling. Ag-RDTs were conducted and interpreted in accordance with the manufacturer’s instructions; results of the BD-Veritor Ag-RDT were determined visually instead of using a Veritor Plus Analyzer.^11^

While molecular testing was used as the reference standard in all three centralised laboratories, the sampling and molecular testing details varied slightly (supplementary material 2). In Breda and Zwolle, OP-N sampling was combined with RT-PCR or transcription-mediated amplification (TMA) testing, respectively. Samples that tested positive by TMA in Zwolle were subsequently tested by RT-PCR to generate a Ct value. The Rotterdam site used combined oropharyngeal and nasopharyngeal (OP-NP) sampling combined with RT-PCR. The platforms used were Roche cobas 6800/8800 (Rotterdam and Breda, respectively) and ABI-7500 (Zwolle) for RT-PCR, and the Hologic Panther system (Aptima SARS-CoV2 assay) for TMA (supplementary material 2).

All staff assessing test results were blinded to the results of the other test. In the first phase of the study, the Ag-RDTs were conducted in accordance with routine test site procedures; participants were therefore informed about the Ag-RDT result but not the subsequent molecular test result. In the second phase of the study, the Ag-RDTs were not conducted according to routine practice and participants were therefore informed about the molecular test result.

In discordant cases (Ag-RDT negative and RT-PCR positive cases) whole genome sequencing (WGS) of the primary clinical sample was performed when the viral load was above the infectiousness cut-off (supplementary material 2).

### Outcomes and statistical analyses

The primary outcomes were the diagnostic accuracies (sensitivity, specificity, positive and negative predictive values with corresponding 95% confidence intervals [CI]) of all three Ag-RDTs and sampling technique combinations, with molecular testing as the reference standard. As the number of individuals without molecular test or Ag-RDT results was very low (n=55 (0.7%); Figure 1), we performed a complete case analysis.

**Figure 1.**
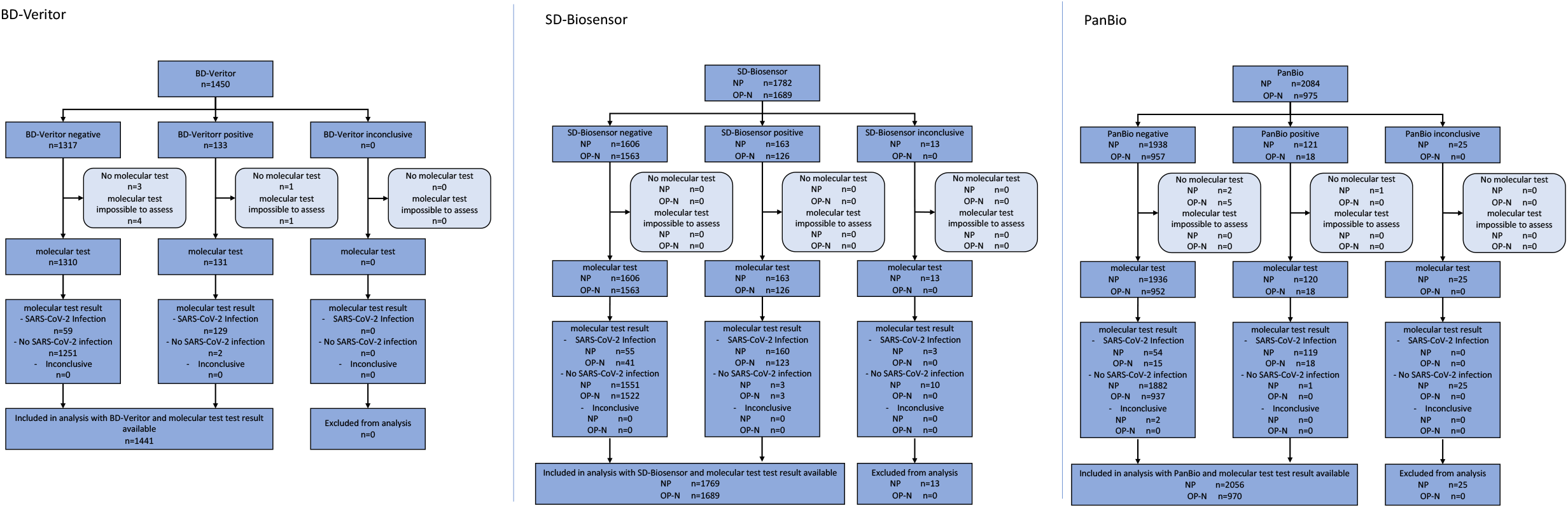
Flow of study participants. BD-Veritor = BD Veritor™ System by Becton Dickinson, SD-Biosensor = Roche/SD Biosensor by Roche Diagnostics, PanBio = PanBio by Abbot.

Secondary outcomes were diagnostic accuracies using a viral load cut-off as a proxy of infectiousness (≥5.2 log10 SARS-CoV-2 E-gene copies/mL), which was the viral load cut-off above which 95% of people with a positive molecular test had a positive virus culture in a recent study by our group^6^, and diagnostic accuracies stratified by presence of symptoms at time of sampling (yes or no), COVID-19 vaccination status (vaccinated with at least one dose yes or no), having had a prior SARS-CoV-2 infection (yes or no), sex (female or male), age (≥16 to ≤40 or >40 to ≤65 or >65), and testing indication (symptoms and/or close contact without symptoms). In an additional analysis, we performed WGS to assess whether false-negative Ag-RDT results could be linked to SARS-CoV-2 variants or specific mutations in the SARS-CoV-2 N-gene (supplementary material 2).

Finally, we used the SARS-CoV-2 test results database of the public health service test sites to identify any missed infections using pseudonymised linkage. Specifically, we determined whether participants who received a negative test result had tested positive in the subsequent 14 days by either molecular test or Ag-RDT, and analysed the interval between the initial and follow-up test. Follow up results of participants were stratified by the results of the initial Ag-RDT and molecular reference tests and by the presence of symptoms at the time of sampling.

### Sample size calculation

Previous diagnostic accuracy studies of Ag-RDTs in people with COVID-19-like symptoms found sensitivities of around 80-85%.^3-5,11,12^ We based our sample size calculation on an expected sensitivity of 80% for each Ag-RDT, with a margin of error of 7%, type I error of 5% and power of 90%. We required approximately 145 positive reference tests for each Ag-RDT-molecular reference test comparison, and per Ag-RDT sampling technique (routinely used versus less invasive). We expected a negligible non-response rate based on previous studies. We anticipated a SARS-CoV-2 prevalence (based on molecular testing) of 10%, and closely monitored molecular test positivity rates over time in order to prolong recruitment as needed.

### Patient and public involvement

Patients and the public were not involved in the design of this research.

## Results

Between 12 April and 14 June 2021, 7980 individuals participated in the study (Figure 1). Results for both a molecular reference test and an Ag-RDT were available for 1441 participants (99.4%) in the BD-Veritor/OP-N sampling group, 1769 participants (99.3%) in the SD-Biosensor/NP sampling group, 1689 participants (100%) in the SD-Biosensor/OP-N sampling group, 2056 participants (98.7%) in the PanBio/NP sampling group, and 970 participants (99.5%) in the PanBio/OP-N sampling group.

The SARS-CoV-2 prevalence in the Netherlands started to decline on 15 May 2021. The required number of positive reference tests had (almost) been reached in Breda and Rotterdam by then. In Zwolle, however, the second phase of the study (PanBio test using less-invasive OP-N sampling) was initiated on 1 June 2021 and was terminated early on 14 June 2021 due to the low PCR test positivity percentage (only 3.4%; 33 positive molecular reference tests in 970 participants). Results of this incomplete evaluation are presented in supplementary Tables S1 and S2 and are not described any further in this manuscript.

The demographic characteristics of the study groups were similar: the mean ages ranged from 37.5 (SD 14.8) to 41.1 years (SD 16.3) and the percentages of female participants between 50.7% and 55.6% (Table 1).

**Table 1.** Baseline characteristics of the study population, stratified by type of rapid antigen test and sampling method.

| Test | BD-Veritor | SD-Biosensor |  | PanBio |
| --- | --- | --- | --- | --- |
| Method of sampling | Routinely used: OP-N | Routinely used: NP | Less invasive: OP-N | Routinely used: NP |
| Inclusion period | 12-30 Apr 2021 | 14-20 Apr 2021 | 3-17 May 2021 | 12-22 Apr 2021 |
| Sample size | N = 1,441 | N = 1,769 | N = 1,689 | N = 2,056 |
| Age [years], mean (SD) <sup>#</sup> | 41.1 (16.3) | 39.5 (15.5) | 37.5 (14.8) | 37.6 (14.8) |
| Sex, female n (%) <sup>†</sup> | 798 (55.6) | 894 (50.7) | 856 (50.8) | 1075 (52.4) |
| Testing indication, n (%) <sup>‡</sup> |  |  |  |  |
| Symptomatic | 501 (34.8) | 952 (53.8) | 759 (44.9) | 1273 (61.9) |
| Pre-/asymptomatic close contact of confirmed SARS-CoV-2 infected individual | 800 (55.6) | 688 (38.9) | 752 (43.9) | 594 (28.9) |
| Other | 73 (5.1) | 92 (5.2) | 93 (5.5) | 91 (4.4) |
| Unknown | 67 (4.6) | 37 (2.1) | 85 (5.0) | 98 (4.8) |
| Vaccinated with at least one dose, n(%) <sup>@</sup> | 152 (10.5) | 96 (5.4) | 224 (13.3) | 167 (8.1) |
| Type of vaccine, n(%) <sup>§</sup> |  |  |  |  |
| Astra Zeneca | 77 (50.7) | 48 (50.0) | 67 (29.9) | 113 (67.7) |
| Janssen |  |  | 7 (3.1) |  |
| Moderna | 7 (4.6) | 5 (5.2) | 19 (8.5) | 9 (5.4) |
| Pfizer | 63 (41.4) | 36 (37.5) | 121 (54.0) | 43 (25.7) |
| Unknown | 5 (3.3) | 7 (7.3) | 10 (4.5) | 2 (1.2) |
| Number of vaccinations received, n(%) <sup>§</sup> |  |  |  |  |
| 1 | 107 (70.4) | 75 (78.1) | 169 (75.4) | 136 (81.4) |
| 2 | 31 (20.4) | 11 (11.5) | 33 (14.7) | 20 (12.0) |
| Unknown | 14 (9.2) | 10 (10.4) | 22 (9.8) | 11 (6.6) |
| At least one prior SARS-CoV-2 infection, n(%) <sup>§</sup> | 102 (7.1) | 187 (10.6) | 196 (11.6) | 134 (6.5) |
| Symptoms at time of sampling, n (%) | 662 (47.2) | 1091 (62.4) | 900 (55.0) | 1470 (74.2) |
| Symptom onset, n (%) <sup>*</sup> |  |  |  |  |
| At day of sampling | 19 (2.9) | 91 (8.3) | 70 (7.8) | 240 (16.3) |
| A day before sampling | 189 (28.5) | 482 (44.2) | 374 (41.6) | 610 (41.5) |
| Two days before sampling | 218 (32.9) | 282 (25.8) | 209 (23.2) | 332 (22.6) |
| Three or more days before sampling | 252 (38.1) | 250 (22.9) | 247 (27.4) | 286 (19.5) |
| Unknown | 15 (2.3) | 15 (1.4) | 19 (2.1) | 20 (1.4) |
| Type of symptoms (self-reported), n (%) <sup>*#</sup> |  |  |  |  |
| Common cold | 570 (86.1) | 948 (86.9) | 768 (85.3) | 1349 (91.8) |
| Shortness of breath | 113 (17.1) | 137 (12.6) | 121 (13.4) | 197 (13.4) |
| Fever | 72 (10.9) | 146 (13.4) | 126 (14.0) | 157 (10.7) |
| Coughing | 308 (46.5) | 450 (41.2) | 342 (38.0) | 584 (39.7) |
| Loss of taste or smell | 24 (3.6) | 43 (3.9) | 41 (4.6) | 55 (3.7) |
| Muscle ache | 88 (13.3) | 137 (12.6) | 100 (11.1) | 143 (9.7) |
| Other symptoms | 37 (5.6) | 18 (1.6) | 54 (6.0) | 74 (5.0) |

NP = deep nasopharyngeal; OP-N = combined oropharyngeal and nasal sampling; SD=standard deviation.
In the Netherlands, individuals are notified of a close contact by the Dutch public health service test-and-trace program, and/or the Dutch contact tracing mobile phone application (the CoronaMelder app) and/or an individual with a confirmed SARS-CoV-2 infection (index case).
<sup>#</sup> Age was not available from 3, 4, 4, and 2 participants in the BD-Veritor group, SD-Biosensor NP group, SD-Biosensor OP-N group, and PanBio NP group, respectively.
<sup>†</sup> Sex not available from 6, 4, 3, and 4 participants in the BD-Veritor group, SD-Biosensor NP group, SD-Biosensor OP-N group, and PanBio NP group, respectively.
<sup>‡</sup> Indication for testing was referral for other reason for 73, 92, 93, and 91 participants and unknown for 67, 37, 85, and 98 participants in the BD-Veritor group, SD-Biosensor NP group, SD-Biosensor OP-N group, and PanBio NP group, respectively.
<sup>@</sup> COVID-19 vaccination status not available from 34, 14, 53, and 72 participants, including 7, 0, 4, and 7 with a positive molecular test result, in the BD-Veritor group, SD-Biosensor NP group, SD-Biosensor OP-N group, and PanBio NP group, respectively.
<sup>§</sup> percentage calculated as proportion of those vaccinated
<sup>§</sup> Previous SARS-CoV-2 infection information not available from 48, 14, 56, and 72 participants in the BD-Veritor group, SD-Biosensor NP group, SD-Biosensor OP-N group, and PanBio NP group, respectively.
<sup>\*</sup> percentage calculated as proportion of those with symptoms at time of sampling
<sup>#</sup> totals add up to a number higher than the number of individuals with symptoms at the time of sampling because individuals could report more than one symptom.

Table 2 shows the results of the primary analysis and the secondary analyses stratified by infectiousness cut-off and presence or absence of symptoms at the time of sampling. Secondary analyses stratified by COVID-19 vaccination status, sex, and age are presented in supplementary Table S3. Sensitivities of all primary and secondary analyses are visualised in Figure 2. Supplementary Tables S4 to S8 show 2×2 tables for each Ag-RDT-sampling technique combination.

**Table 2.** Diagnostic accuracy variables of three rapid antigen tests, with different sampling methods. Values are percentages (95% confidence interval) unless stated otherwise.

| Analysis | Sampling method | No. | Prevalence* (%) | Sensitivity | Specificity | PPV | NPV |
| --- | --- | --- | --- | --- | --- | --- | --- |
| **BD-Veritor System (Beckton Dickinson)** |  |  |  |  |  |  |  |
| Primary analysis | OP-N | 1441 | 13.0 | 68.6<br>(61.5 to 75.2) | 99.8<br>(99.4 to 100.0) | 98.5<br>(94.6 to 99.8) | 95.5<br>(94.2 to 96.6) |
| Secondary (stratified) analysis:<br>Infectiousness viral load cut-off¶ | OP-N | 1441 | 10.1 | 85.6<br>(78.9 to 90.9) | 99.5<br>(99.0 to 99.8) | 95.4<br>(90.3 to 98.3) | 98.4<br>(97.6 to 99.0) |
| Symptoms present at sampling‡:<br>Yes | OP-N | 662 | 18.1 | 75.8<br>(67.2 to 83.2) | 99.8<br>(99.0 to 100.0) | 98.9<br>(94.1 to 100.0) | 94.9<br>(92.8 to 96.6) |
| No | OP-N | 742 | 8.0 | 55.9<br>(42.4 to 68.8) | 99.9<br>(99.2 to 100.0) | 97.1<br>(84.7 to 99.9) | 96.3<br>(94.7 to 97.6) |
| **SD-Biosensor (Roche Diagnostics)** |  |  |  |  |  |  |  |
| Primary analysis | NP | 1769 | 12.2 | 74.4<br>(68.0 to 80.1) | 99.8<br>(99.4 to 100.0) | 98.2<br>(94.7 to 99.6) | 96.6<br>(95.6 to 97.4) |
|  | OP-N | 1689 | 9.7 | 75.0<br>(67.7 to 81.4) | 99.8<br>(99.4 to 100.0) | 97.6<br>(93.2 to 99.5) | 97.4<br>(96.5 to 98.1) |
| Secondary (stratified) analysis:<br>Infectiousness viral load cut-off¶ | NP | 1769 | 10.3 | 87.9<br>(82.3 to 92.3) | 99.8<br>(99.4 to 100.0) | 98.2<br>(94.7 to 99.6) | 98.6<br>(97.9 to 99.1) |
|  | OP-N | 1689 | 8.3 | 83.7<br>(76.5 to 89.4) | 99.5<br>(99.0 to 99.8) | 93.7<br>(87.9 to 97.2) | 98.5<br>(97.8 to 99.1) |
| Symptoms present at sampling‡:<br>Yes | NP | 1091 | 13.8 | 83.4<br>(76.5 to 89.0) | 99.8<br>(99.2 to 100.0) | 98.4<br>(94.5 to 99.8) | 97.4<br>(96.2 to 98.3) |
|  | OP-N | 900 | 12.7 | 78.9<br>(70.3 to 86.0) | 99.7<br>(99.1 to 100.0) | 97.8<br>(92.4 to 99.7) | 97.0<br>(95.6 to 98.1) |
| No | NP | 658 | 9.6 | 54.0<br>(40.9 to 66.6) | 99.8<br>(99.1 to 100.0) | 97.1<br>(85.1 to 99.9) | 95.3<br>(93.4 to 96.9) |
|  | OP-N | 735 | 6.3 | 63.0<br>(47.5 to 76.8) | 100.0<br>(99.5 to 100.0) | 100.0<br>(88.1 to 100.0) | 97.6<br>(96.2 to 98.6) |
| **PanBio (Abbott)** |  |  |  |  |  |  |  |
| Primary analysis | NP | 2056 | 8.4 | 68.8<br>(61.3 to 75.6) | 99.9<br>(99.7 to 100.0) | 99.2<br>(95.4 to 100.0) | 97.2<br>(96.4 to 97.9) |
| Secondary (stratified) analysis:<br>Infectiousness viral load cut-off¶§ | NP | 2039 | 5.9 | 89.3<br>(82.3 to 94.2) | 99.9<br>(99.6 to 100.0) | 98.2<br>(93.6 to 99.8) | 99.3<br>(98.9 to 99.6) |
| Symptoms present at sampling‡:<br>Yes | NP | 1470 | 9.0 | 72.2<br>(63.7 to 79.6) | 99.9<br>(99.6 to 100.0) | 99.0<br>(94.4 to 100.0) | 97.3<br>(96.3 to 98.1) |
| No | NP | 511 | 6.7 | 55.9<br>(37.9 to 72.8) | 100.0<br>(99.2 to 100.0) | 100.0<br>(82.4 to 100.0) | 97.0<br>(95.0 to 98.3) |

NC = not calculated because all Ag-RDT results were negative; NP = deep nasopharyngeal; OP-N = combined oropharyngeal and nasal sampling; PPV=positive predictive value; NPV=negative predictive value.
\*SARS-CoV-2 infection based on molecular test result.
‡Symptoms not available for 37, 20, 53, and 75 participants, including 9, 1, 4, and 7 with a positive molecular test result, in the BD-Veritor group, SD-Biosensor NP group, SD-Biosensor OP-N group, and PanBio NP group, respectively.
¶Viral load cut-off for infectiousness, defined as viral load above which 95% of people with a positive RT-PCR test result had a positive viral culture<sup>6</sup>, was 5.2 log<sub>10</sub> SARS-CoV-2 E-gene copies/mL.
§Viral load unavailable for 17 participants in the PanBio NP group.

**Figure 2.**
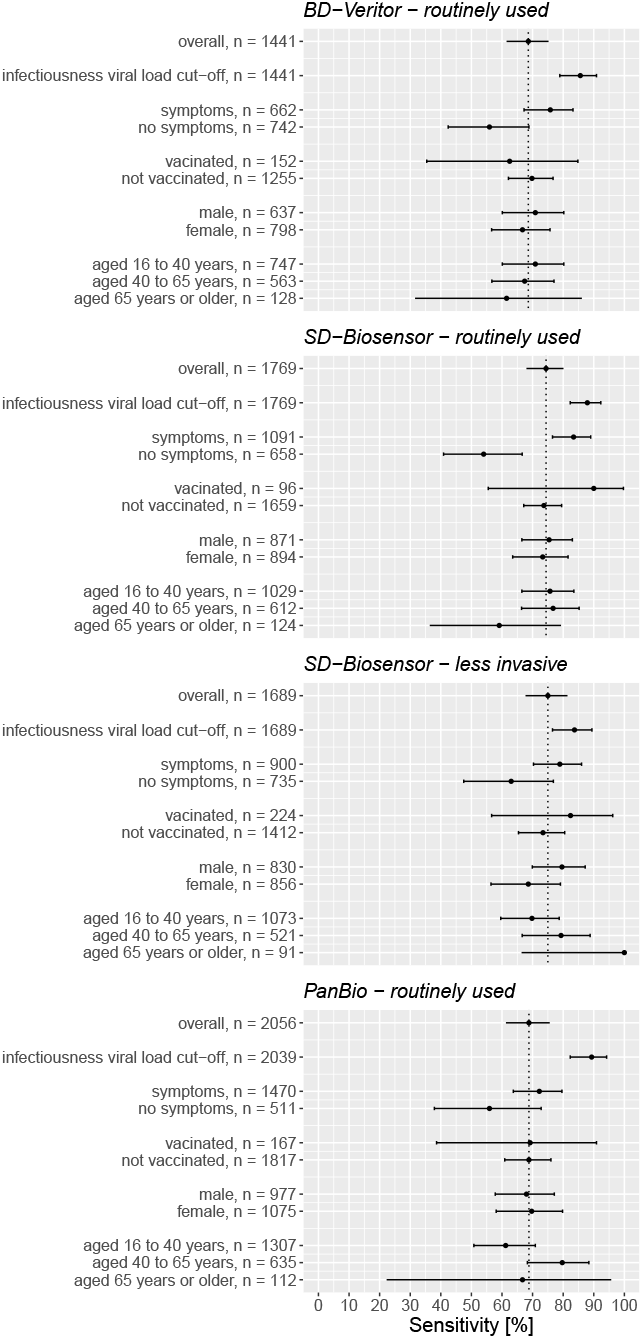
Sensitivities with 95% confidence intervals of the various antigen rapid test-moleculare reference standard test comparisons, stratified according to symptomatology, COVID-19 vaccination status, sex and age. BD-Veritor = BD Veritor™ System by Becton Dickinson, SD-Biosensor = Roche/SD Biosensor by Roche Diagnostics, PanBio = PanBio by Abbot.

### Routinely used Ag-RDT sampling method

SARS-CoV-2 prevalence (by molecular reference test) was 13.0% (188/1441) in the BD-Veritor group, 12.2% (215/1769) in the SD-Biosensor group, and 8.4% (173/2056) in the PanBio group. Overall sensitivities were 68.6% [61.5%-75.2%] for BD-Veritor, 74.4% [68.0%-80.1%] for SD-Biosensor, and 68.8% [61.3%-75.6%] for PanBio (Table 2, Figure 2).

Among those with a positive molecular test result, the percentage of participants with a viral load above the cut-off as a proxy for infectiousness was 77.7% (146/188) in the BD-Veritor group, 85.1% (183/215) in the SD-Biosensor group, and 70.0% (121/173) in the PanBio group. Using this viral load cut-off, the sensitivities were 85.6% [78.9%-90.9%] for BD-Veritor, 87.9% [82.3%-92.3%] for SD-Biosensor, and 89.3% [82.3%-94.2%] for PanBio.

Sensitivities ranged from 72.2% to 83.4% in individuals who were symptomatic at the time of sampling and from 54.0% to 55.9% in those who were asymptomatic (Table 2, Figure 2). We found no evidence of a differential impact on diagnostic accuracy by COVID-19 vaccination status, sex, and age (Figure 2, supplementary Table S3).

Specificities were >99%, and positive and negative predictive values were >95%, for all three Ag-RDT in most analyses (Table 2 and supplementary Table S3).

### Less invasive OP-N sampling method combined with SD-Biosensor

The SARS-CoV-2 prevalence (by molecular reference test) was 9.7% (164/1689) and the sensitivity was 75.0% [67.7%-81.4%] (Table 2, Figure 2).

Among those with a positive molecular test result, the percentage of participants with a viral load above the cut-off as a proxy for infectiousness was 86.0% (141/164). Using the viral load cut-off as a proxy for infectiousness, the sensitivity was 83.7% [76.5%-89.4%].

Sensitivities were 78.9% in symptomatic individuals and 63.0% in those who were asymptomatic at the time of sampling (Table 2, Figure 2). We found no evidence of a differential impact on diagnostic accuracy by COVID-19 vaccination status, sex, and age (supplementary table S3).

Specificities were >99%, and positive and negative predictive values were >95%, in most analyses (Table 2 and supplementary Table S3).

### WGS results of discordant pairs

In total 209 samples had discordant results, of which 79 samples had viral loads above the infectiousness cut-off. Nine samples could not be retrieved for WGS, 23 samples were excluded due to a negative result or high Ct values (>32) in the confirmatory RT-PCR prior to WGS, and WGS was not successful in 12 samples. Of the remaining 35 samples, two samples had a reference coverage <70%, 10 samples of 70-90%, and 23 samples of >90%. The latter 23 samples were submitted to GISAID (supplementary Table S9): 22 were typed as B.1.1.7 (Alpha, V1) and one as B.1.351 (Beta, V2). A complete N-gene sequence was retrieved from all 33 samples with a coverage ≥70%. No N-gene aminoacid changes were found in 22 samples according to their respective lineage (either Alpha or Beta). In the remaining 11 samples (Alpha variants), the following single amino acid changes were found respective to their lineage: D63Y, G204P (six times), P151L, M234I (two times) and Q303R.

### Follow-up

All but six participants could be linked with the national test results database of the public health services for the follow up analyses. In the first phase of the study, a negative Ag-RDT result was communicated to 4847 participants, of which 168 (3.5%) were false negative based on a positive molecular test. At least one subsequent test (Ag-RDT or molecular test) was registered in the national test results database within 14 days after study participation for 887/4847 (18.3%) participants: for 103/168 (61.3%) of participants with a false negative Ag-RDT and for 786/4679 (16.8%) of participants with a true negative Ag-RDT (χ^2^ =215, p<.001). Overall, 213/4847 (4.4%) Ag-RDT negative participants tested SARS-CoV-2 positive within 14 days after their negative Ag-RDT result in our study: 55.4% (93/168) of false negatives and 2.6% (120/4679) of true negatives (χ^2^ =1076, p<.001). The positivity percentages were 90.3% (93/103) and 15.3% (120/786) for the false negative and true negative Ag-RDT groups, respectively (χ^2^ =281, p<.001). In the second phase, in which the molecular test result was communicated, 1.1% (28/2461) of participants with a negative molecular test had a positive test registered in the national test results database within the 14 day follow up period.

In the first phase, the interval between a negative Ag-RDT in our study and a positive test in the 14 day follow-up period was shorter for false negatives (median 3 days, interquartile range (IQR) 3 days) than for true negatives (median 5 days, IQR 3 days; U=2810, p<.001). In the second phase, the median interval between a negative molecular test and a positive follow-up test was 6 days (IQR 5 days).

In the first phase of the study, being asymptomatic at the time of sampling was associated with a higher likelihood of testing positive during the 14 day follow-up period. Four percent (70/1752) of participants with a true negative Ag-RDT without symptoms, tested positive during follow up compared to 1.6% (45/2802) of those with symptoms (χ^2^ =25, p<.001). These percentages were 62.9% (44/70) and 46.2% (42/91), respectively, for participants with a false negative Ag-RDT (χ^2^ =4.4, p=.035). In phase 2, similar relative differences were found between participants who were asymptomatic and symptomatic at time of sampling but was found not to be statistically significant: 1.7% (16/965) of participants with a negative molecular test and no symptoms at initial testing tested positive versus 0.8% (12/1427) of those with symptoms (χ^2^ =3.3, p=.068).

## Discussion

The BD-Veritor, SD-Biosensor and PanBio lateral flow Ag-RDTs are the three most used SARS-CoV-2 point-of-care tests in the Netherlands. They underwent limited diagnostic accuracy evaluations prior to their approval for use in the public testing programme but were never evaluated in a large community-based study with nationwide reach. In addition, the public’s desire to move away from deep NP sampling increased over time, but the diagnostic accuracies of the SD-Biosensor and PanBio tests using OP-N sampling were not yet known.

Our study found that the three Ag-RDTs combined with their routine sampling techniques had sensitivities of 68.6% to 74.4%, increasing to at least 85.6% after a viral load cut-off was applied as a proxy for infectiousness. Sensitivities ranged from 72.2% to 83.4% in individuals who were symptomatic at the time of sampling and from 54.0% to 55.9% in those who were asymptomatic. We found no evidence of a differential impact on diagnostic accuracy of COVID-19 vaccination status, sex, and age. For SD-Biosensor, the less invasive OP-N sampling technique yielded comparable sensitivities in the primary and secondary stratified analyses as the deep NP approach. Specificities and positive and negative predictive values were high for all Ag-RDT sampling technique combinations.

Follow-up analyses of persons with false negative Ag-RDT results show that more than 55% (symptomatic vs asymptomatic at time of initial sampling: 46% vs 63%) tested positive in the 14 days after the initial test, whereas positive test results within 14 days after initial testing occurred in 1.1% of people with a negative initial molecular test.

### Comparison with other studies

The overall unstratified sensitivities of the Ag-RDTs in our study were substantially lower than those reported in Ag-RDT evaluations performed earlier in the pandemic in the Netherlands.^11,12^ We hypothesize that the reason for this is because only symptomatic individuals could access SARS-CoV-2 testing in the Netherlands until 1 December 2020. The sensitivities in our study for individuals who were symptomatic at the time of sampling were indeed similar to those in the earlier evaluation studies. The sensitivities in our study for individuals who were asymptomatic at the time of sampling are in line with those that we found in our recent study in asymptomatic and presymptomatic close contacts.^6^ The positivity percentages during the 14-day follow-up period of people with a negative initial molecular test in our study was 1.1%, which was slightly lower than the 1.7% found in our previous study among close contacts. This can likely be attributed to the fact that close contacts are at higher risk of testing SARS-CoV-2-positive than those testing for other reasons.^6^

### Strengths and limitations of this study

Strengths include the protocolised nature of the study, the large sample size covering multiple test sites nationwide, the high data completeness, collection of samples for the index and reference tests at the same time, the implementation of index and reference tests by trained staff who were blinded to the result of the other test, and the availability of follow up information for participants who received negative test results.

Our study also has some limitations. First, the reference standards that we used were molecular tests, but platforms and test kits used differed among the three centralised laboratories. However, the diagnostic accuracies of all molecular tests used are similarly high^13,14^, and we therefore believe that this has not influenced our findings significantly. In addition, Ct values determined by the different platforms were comparable (supplementary material 2). Second, we were unable to meet the predefined sample size in the PanBio/OP-N sampling group. These results are therefore not sufficiently robust and should be interpreted with great caution. Third, while the Delta variant is the dominant SARS-CoV-2 variant in the Netherlands at time of writing, its prevalence was only around 8.6% during the last week of inclusions. However, we checked whether false-negative Ag-RDT results could be linked to specific virus lineages by WGS and we did not find a signal confirming this hypothesis.

Fourth, we relied on infectiousness viral load cut-offs that were determined in our previous study in an almost completely unvaccinated population, whereas the proportion of vaccinated individuals in the present study reached 20% at the end of the study. Whether this would have impacted the applied viral load cut-offs is unknown. Fifth, our sample size calculation was based on the primary analysis and the diagnostic accuracy parameters are therefore less precise for the secondary stratified analyses. For example, a differential impact of COVID-19 vaccination status on diagnostic accuracy of Ag-RDTs might be anticipated given that vaccinated individuals have shorter periods of high viral loads and lower virus viability by Ct values than unvaccinated individuals, resulting in lower transmissibility^15-19^ Further studies on the potential impact of COVID-19 vaccination on Ag-RDT diagnostic accuracies are warranted. Finally, we did not actively follow-up participants who had received a negative test result but collected follow-up information from the public health services test result database through pseudonymised linkage. Active follow-up, including repeat testing in all study participants, would have reduced the uncertainty around false negative Ag-RDT results completely, as was also recommended in a recent guidance paper.^20^ Unfortunately, we could not implement this because of ethical and logistic constraints, as our study was embedded in busy public health service test sites. Also, we cannot be certain that all positive tests within the maximum incubation period after a negative initial test represent false negative tests; they could also have resulted from a new SARS-CoV-2 exposure after the initial test. However, this is true for individuals with a negative Ag-RDT and for those with a negative molecular test at inclusion, and the difference in positivity percentages during follow-up was strikingly large.

### Policy implications

The advantages of Ag-RDTs compared with molecular testing are simplified logistics and reduced delays. The more frequent use of Ag-RDTs instead of molecular testing will inevitably lead to an increase in the number of missed infections, especially when used in asymptomatic individuals, e.g., for travelling and/or access to events. We observed that about 60% of persons with false negative Ag-RDT results returned for testing, and 55% tested positive, within 14 days after the initial negative test with a median delay of 3 days. About half of them did not have symptoms at the time of initial sampling, and those participants were more likely to have a positive follow-up test (symptomatic vs asymptomatic at time of initial sampling: 46% vs 63%). This suggests that a considerable portion of individuals adhered to the advice to return for testing if symptoms develop or worsen after a negative test and also illustrates the importance of emphasizing the need for re-testing to individuals with a negative Ag-RDT should such events occur. However, as over half of all infections are estimated to occur before the onset of symptoms, these missed early infections do pose a relevant risk for further transmission.^21,22^ Furthermore, the 45% of individuals with a false negative Ag-RDT who are never diagnosed pose an even greater risk of onward transmission. The extent to which the advantages of Ag-RDTs outweigh their lower sensitivities depends on several aspects, including the potential consequences of missed infections.

Furthermore, as the prevalence of SARS-CoV-2 declines, the positive predictive value of Ag-RDT results will decrease, meaning a larger proportion of positive test results will be false positive.^3^ In such circumstances, the risk of false positive results with Ag-RDTs could be mitigated by confirmatory molecular testing.

### Conclusions

Compared to molecular testing, the sensitivities of three widely used SARS-CoV-2 Ag-RDTs when applying the routinely used sampling techniques were at least 68% and increased to at least 85% after a viral load cut-off was applied as a proxy for infectiousness. Sensitivities ranged between 54.0% and 55.9% in those who were asymptomatic at time of sampling, meaning that around half of infections might be missed in this population. Follow-up analyses revealed that over 55% of asymptomatic persons with a false negative Ag-RDT result tested positive within 14 days after the initial test emphasizing the need for re-testing should symptoms develop and education of the public about a high potential of false negative Ag-RDTs when asymptomatic. In high-risk situations, such as testing of vulnerable people in care facilities, severely ill patients, or healthcare workers, molecular testing remains the preferred option. For SD-Biosensor, the less invasive OP-N sampling technique yielded comparable diagnostic accuracies for Ag-RDT-molecular test comparisons as the deep NP approach. Adopting this more convenient sampling method might reduce the threshold for SARS-CoV-2 testing.

## Supporting information

STARD-checklist

## Data Availability

All data produced in the present study are available upon reasonable request to the authors

## Acknowledgements

We thank the participants, and study staff at the participating public health service test sites, participating laboratories, the University Medical Center Utrecht, and RIVM for their contributions to the study. A special thanks to Esther Stiefelhagen, Roel Ensing, Wendy Mouthaan, Lieke Brouwer, and Timo Boelsums. Written permission was obtained from all five of them to list their names. ES, RE, WH, LB, and TB did not receive any compensation for their contributions.

## Contributors

KGMM initiated the study. RPV, IKV, KGMM, WvdB, EL, RM, CRSN-I, LH, JHHMvdW, SvdH, and ES designed the study. RPV, IKV, CRSN-I and ES coordinated the study. WvdB, SDP, RM, ZI, COdS, SBD, and MJB were responsible for laboratory analyses and data processing. RPV, IKV and ES verified the underlying data. ES performed the statistical analysis in close collaboration with RPV, IKV and KGMM. RPV, IKV, KGMM, WvdB, JHHMvdW and ES drafted the first version of the manuscript. All authors critically read the manuscript and provided feedback. All authors approved the submission of the current version of the manuscript. The corresponding author attests that all listed authors meet authorship criteria and that no others meeting the criteria have been omitted.

## Ethical approval

Not required because the study was judged by the METC Utrecht to be outside the scope of the Dutch Medical Research Involving Human Subjects Act (protocol No 21-146/C). All participants signed an informed consent form before any study procedure.

## Data sharing

Individual participant data collected during the study will be available, after deidentification of all participants. Data will be available to researchers who provide a methodologically sound proposal to achieve the aims in the approved proposal. Proposals should be directed to the corresponding author to gain access to the data. Data requestors will need to sign a data sharing agreement. The corresponding author (RPV, the manuscript’s guarantor) affirms that the manuscript is an honest, accurate, and transparent account of the study being reported; that no important aspects of the study have been omitted; and that any discrepancies from the study as originally planned (and, if relevant, registered) have been explained.

## Dissemination to participants and related patient and public communities

Our study findings will be disseminated to the the Dutch Outbreak Management Team that provides guidance to the Ministry of Health, Welfare, and Sport on policy regarding COVID-19. If any changes to the nationwide testing policy will be made, findings will be automatically disseminated to the wider public.

## Additional information (containing supplementary information line (if any) and corresponding author line)

The study protocol is available upon request by contacting Roderick Venekamp at.

