## supplementary appendix for "Diagnostic accuracy of three prevailing rapid antigen tests for detection of SARS-CoV-2 infection in the general population: cross sectional study"

### Table of contents

**Table S1** Baseline characteristics of the study population included in the PanBio test with less invasive sampling technique evaluation. 2

**Table S2** Diagnostic accuracy variables of PanBio-molecular reference standard test comparison with less invasive sampling. Values are percentages (95% confidence interval) unless stated otherwise. 3

**Table S3** Diagnostic accuracy variables of additional secondary analyses of three rapid antigen tests, with different sampling methods. Values are percentages (95% confidence interval) unless stated otherwise. 4

**Table S4** Two-by-two tables used in primary and secondary analysis to determine diagnostic accuracy parameters of the *BD Veritor™ System by Becton Dickinson* ('BD-Veritor') using routine sampling (OP-N) 6

**Table S5** Two-by-two tables used in primary and secondary analysis to determine diagnostic accuracy parameters of the *Roche/SD Biosensor by Roche Diagnostics* ('SD-Biosensor') using routine sampling (NP) 8

**Table S6** Two-by-two tables used in primary and secondary analysis to determine diagnostic accuracy parameters of the *Roche/SD Biosensor by Roche Diagnostics* ('SD-Biosensor') using less invasive sampling (OP-N) 10

**Table S7** Two-by-two tables used in primary and secondary analysis to determine diagnostic accuracy parameters of the *PanBio by Abbot* ('PanBio') using routine sampling (NP) 12

**Table S8** Two-by-two tables used in primary and secondary analysis to determine diagnostic accuracy parameters of the *PanBio by Abbot* ('PanBio') using less invasive sampling (OP-N) 14

**Table S9** GISAID data contributors acknowledgement table 16

**Supplementary Material 1:** short questionnaire (translated from Dutch) 17

**Supplementary Material 2:** Specimen collection, SARS-CoV-2 diagnostic testing, and SARS-CoV-2 virus culture procedures 18

**References** 21

**Table S1** Baseline characteristics of the study population included in the PanBio test with less invasive sampling technique evaluation.

| Test | PanBio |
|---|---|
| Method of sampling | Less invasive: OP-N |
| Inclusion period | 1-14 Jun 2021 |
| Sample size | N = 970 |
| Age [years], mean (SD)[#] | 38.4 (14.9) |
| Sex, female n (%)[†] | 473 (49.1) |
| Testing indication, n (%)[!] | |
| Symptomatic | 613 (63.2) |
| Pre-/asymptomatic close contact of confirmed SARS-CoV-2 infected individual | 236 (24.3) |
| Other | 90 (9.3) |
| Unknown | 31 (3.2) |
| Vaccinated with at least one dose, n(%)[@] | 301 (31.0) |
| Type of vaccine, n(%)[$] | |
| Astra Zeneca | 71 (23.6) |
| Janssen | 20 (6.6) |
| Moderna | 23 (7.6) |
| Pfizer | 182 (60.5) |
| Unknown | 6 (2.0) |
| Number of vaccinations received, n(%)[$] | |
| 1 | 238 (79.1) |
| 2 | 53 (17.6) |
| Unknown | 11 (3.7) |
| At least one prior SARS-CoV-2 infection, n(%)[§] | 57 (5.9) |
| Symptoms at time of sampling, n (%)[‡] | 664 (69.7) |
| Symptom onset, n (%)* | |
| At day of sampling | 56 (8.4) |
| A day before sampling | 306 (46.1) |
| Two days before sampling | 172 (25.9) |
| Three or more days before sampling | 139 (20.9) |
| Unknown | 3 (0.5) |
| Type of symptoms (self-reported), n (%)*[#] | |
| Common cold | 610 (91.9) |
| Shortness of breath | 82 (12.3) |
| Fever | 92 (13.9) |
| Coughing | 302 (45.5) |
| Loss of taste or smell | 28 (4.2) |
| Muscle ache | 44 (6.6) |
| Other symptoms | 17 (2.6) |

OP-N = combined oropharyngeal and nasal sampling; SD=standard deviation.
In the Netherlands, individuals are notified of a close contact by the Dutch public health service test-and-trace program, and/or the Dutch contact tracing mobile phone application (the CoronaMelder app) and/or an individual with a confirmed SARS-CoV-2 infection (index case).
[#] Age was not available from 6 participants.
[†] Sex not available from 6 participants.
[!] Indication for testing was referral for other reason for 90 participants and unknown for 31 participants.
[@] COVID-19 vaccination status not available from 17 participants, including 2 with a positive molecular test result.
[$] percentage calculated as proportion of those vaccinated
[§] Previous SARS-CoV-2 infection information not available from 20 participants.
‡Symptoms not available for 18 participants, including 2 with a positive molecular test result.
* percentage calculated as proportion of those with symptoms at time of sampling
[#] totals add up to a number higher than the number of individuals with symptoms at the time of sampling because individuals could report more than one symptom.

**Table S2** Diagnostic accuracy variables of PanBio-molecular reference standard test comparison with less invasive sampling. Values are percentages (95% confidence interval) unless stated otherwise.

| Analysis | Sampling | No. | Prevalence* (%) | Sensitivity | Specificity | PPV | NPV |
|---|---|---|---|---|---|---|---|
| **PanBio (Abbott)** | | | | | | | |
| Primary analysis | OP-N | 970 | 3.4 | 54.5 (36.4 to 71.9) | 100.0 (99.6 to 100.0) | 100.0 (81.5 to 100.0) | 98.4 (97.4 to 99.1) |
| Secondary (stratified) analysis: | | | | | | | |
| Infectiousness viral load cut-off¶ | OP-N | 970 | 2.8 | 66.7 (46.0 to 83.5) | 100.0 (99.6 to 100.0) | 100.0 (81.5 to 100.0) | 99.1 (98.2 to 99.6) |
| Symptoms present at sampling‡: | | | | | | | |
| Yes | OP-N | 664 | 3.0 | 65.0 (40.8 to 84.6) | 100.0 (99.4 to 100.0) | 100.0 (75.3 to 100.0) | 98.9 (97.8 to 99.6) |
| No | OP-N | 288 | 3.8 | 27.3 (6.0 to 61.0) | 100.0 (99.4 to 100.0) | 100.0 (75.3 to 100.0) | 98.9 (97.8 to 99.6) |
| Vaccinated (at least 1 vaccination)[@]: | | | | | | | |
| Yes | OP-N | 301 | 2.0 | 50.0 (11.8 to 88.2) | 100.0 (98.8 to 100.0) | 100.0 (29.2 to 100.0) | 99.0 (97.1 to 99.8) |
| No | OP-N | 652 | 3.8 | 52.0 (31.3 to 72.2) | 100.0 (99.4 to 100.0) | 100.0 (75.3 to 100.0) | 98.1 (96.7 to 99.0) |
| Previous SARS-CoV-2 infection[§]: | | | | | | | |
| Yes | OP-N | 57 | 7.0 | 0.0 (0.0 to 60.2) | 100.0 (93.3 to 100.0) | NC1 | 93.0 (83.0 to 98.1) |
| No | OP-N | 893 | 2.9 | 57.7 (36.9 to 76.6) | 100.0 (99.6 to 100.0) | 100.0 (78.2 to 100.0) | 98.7 (97.8 to 99.4) |
| Sex[†]: | | | | | | | |
| Female | OP-N | 473 | 3.2 | 40.0 (16.3 to 67.7) | 100 (99.2 to 100) | 100 (54.1 to 100) | 98.1 (96.4 to 99.1) |
| Male | OP-N | 491 | 3.7 | 66.7 (41.0 to 86.7) | 100 (99.2 to 100) | 100 (73.5 to 100) | 98.7 (97.3 to 99.5) |
| Age [years][#]: | | | | | | | |
| ≥16 to ≤40 | OP-N | 593 | 4.2 | 48.0 (27.8 to 68.7) | 100 (99.4 to 100) | 100 (73.5 to 100) | 97.8 (96.2 to 98.8) |
| >40 to ≤65 | OP-N | 317 | 2.5 | 75.0 (34.9 to 96.8) | 100 (98.8 to 100) | 100 (54.1 to 100) | 99.4 (97.7 to 99.9) |
| >65 | OP-N | 54 | 0.0 | NC2 | NC2 | NC2 | NC2 |
| Testing indication[!]: | | | | | | | |
| symptomatic | OP-N | 613 | 2.6 | 75.0 (47.6 to 92.7) | 100 (99.4 to 100) | 100 (73.5 to 100) | 99.3 (98.3 to 99.8) |
| close contact with SARS-CoV-2 infected individual | OP-N | 236 | 5.1 | 25.0 (5.5 to 57.2) | 100 (98.4 to 100) | 100 (29.2 to 100) | 96.1 (92.8 to 98.2) |

PPV=positive predictive value; NPV=negative predictive value; OP-N = combined oropharyngeal and nasal sampling; NC1 = not calculated because all Ag-RDT results were negative; NC2 = not calculated because all molecular test results were negative.
*SARS-CoV-2 infection based on molecular test result.
¶Viral load cut-off for infectiousness, defined as viral load above which 95% of people with a positive RT-PCR test result had a positive viral culture[1] was 5.2 log10 SARS-CoV-2 E-gene copies/mL.
‡Symptoms not available for 18 participants, including 2 with a positive molecular test result.
[@] COVID-19 vaccination status not available from 17 participants, including 2 with a positive molecular test result.
[§] Previous SARS-CoV-2 infection information not available from 20 participants.
[†] Sex not available from 6 participants.
[#] Age was not available from 6 participants.
[!] Indication for testing was referral for other reason for 90 participants and unknown for 31 participants.

**Table S3** Diagnostic accuracy variables of additional secondary analyses of three rapid antigen tests, with different sampling methods. Values are percentages (95% confidence interval) unless stated otherwise.

| Analysis | Sampling | No. | Prevalence* (%) | Sensitivity | Specificity | PPV | NPV |
|---|---|---|---|---|---|---|---|
| **BD-Veritor System (Beckton Dickinson), n (OP-N) = 1441** | | | | | | | |
| Vaccinated (at least 1 vaccination)[@]: | | | | | | | |
| Yes | OP-N | 152 | 10.5 | 62.5 (35.4 to 84.8) | 100.0 (97.3 to 100.0) | 100.0 (69.2 to 100.0) | 95.8 (91.0 to 98.4) |
| No | OP-N | 1255 | 12.9 | 69.8 (62.1 to 76.7) | 99.8 (99.3 to 100.0) | 98.3 (93.9 to 99.8) | 95.7 (94.4 to 96.8) |
| Previous SARS-CoV-2 infection[§]: | | | | | | | |
| Yes | OP-N | 102 | 10.8 | 64.6 (30.8 to 89.1) | 100 (96.0 to 100) | 100 (59.0 to 100) | 95.8 (89.6 to 98.8) |
| No | OP-N | 1291 | 12.7 | 70.1 (62.5 to 77.0) | 99.8 (99.4 to 100) | 98.3 (94.0 to 99.8) | 95.8 (94.5 to 96.9) |
| Sex[†]: | | | | | | | |
| Female | OP-N | 798 | 12.8 | 66.7 (56.6 to 75.7) | 99.9 (99.2 to 100) | 98.6 (92.2 to 100) | 95.3 (93.5 to 96.7) |
| Male | OP-N | 637 | 13.5 | 70.9 (60.1 to 80.2) | 99.8 (99.0 to 100) | 98.4 (91.3 to 100) | 95.7 (93.6 to 97.2) |
| Age [years] [#]: | | | | | | | |
| ≥16 to ≤40 | OP-N | 747 | 11.5 | 70.9 (60.1 to 80.2) | 100 (99.4 to 100) | 100 (94.1 to 100) | 96.4 (94.7 to 97.6) |
| >40 to ≤65 | OP-N | 563 | 15.8 | 67.4 (56.7 to 77.0) | 100 (99.2 to 100) | 100 (94.0 to 100) | 94.2 (91.8 to 96.1) |
| >65 | OP-N | 128 | 10.2 | 61.5 (31.6 to 86.1) | 98.3 (93.9 to 99.8) | 80.0 (44.4 to 97.5) | 95.8 (90.4 to 98.6) |
| Testing indication[¦]: | | | | | | | |
| symptomatic | OP-N | 501 | 16.0 | 72.5 (61.4 to 81.9) | 99.5 (98.3 to 99.9) | 96.7 (88.5 to 99.6) | 95.0 (92.5 to 96.8) |
| close contact with SARS-CoV-2 infected individual | OP-N | 800 | 11.6 | 68.8 (58.4 to 78.0) | 100 (99.5 to 100) | 100 (94.4 to 100) | 96.1 (94.4 to 97.3) |
| **SD-Biosensor (Roche Diagnostics), n (NP) = 1769, n (OP-N) = 1689** | | | | | | | |
| Vaccinated (at least 1 vaccination)[@]: | | | | | | | |
| Yes | NP | 96 | 10.4 | 90.0 (55.5 to 99.7) | 100.0 (95.8 to 100.0) | 100.0 (66.4 to 100.0) | 98.9 (93.8 to 100.0) |
| | OP-N | 224 | 7.6 | 82.4 (56.6 to 96.2) | 100.0 (98.2 to 100.0) | 100.0 (76.8 to 100.0) | 98.6 (95.9 to 99.7) |
| No | NP | 1659 | 12.4 | 73.7 (67.1 to 79.5) | 99.8 (99.4 to 100.0) | 98.1 (94.4 to 99.6) | 96.4 (95.3 to 97.3) |
| | OP-N | 1412 | 10.1 | 73.4 (65.4 to 80.5) | 99.8 (99.4 to 100.0) | 98.1 (93.4 to 99.8) | 97.1 (96.0 to 97.9) |
| Previous SARS-CoV-2 infection[§]: | | | | | | | |
| Yes | NP | 187 | 10.2 | 68.4 (43.4 to 87.4) | 100.0 (97.8 to 100.0) | 100.0 (75.3 to 100.0) | 96.6 (92.6 to 98.7) |
| | OP-N | 196 | 5.6 | 54.5 (23.4 to 83.3) | 99.5 (97.0 to 100.0) | 85.7 (42.1 to 99.6) | 97.4 (93.9 to 99.1) |
| No | NP | 1568 | 12.5 | 75.0 (68.3 to 80.9) | 99.8 (99.4 to 100.0) | 98.0 (94.3 to 99.6) | 96.5 (95.5 to 97.4) |
| | OP-N | 1437 | 10.4 | 75.8 (68.2 to 82.5) | 99.9 (99.6 to 100.0) | 99.1 (95.2 to 100.0) | 97.3 (96.3 to 98.1) |
| Sex[†]: | | | | | | | |
| Female | NP | 894 | 11.3 | 73.3 (63.5 to 81.6) | 99.6 (98.9 to 99.9) | 96.1 (89.0 to 99.2) | 96.7 (95.2 to 97.8) |
| | OP-N | 856 | 8.2 | 68.6 (56.4 to 79.1) | 99.9 (99.3 to 100) | 98.0 (89.1 to 99.9) | 97.3 (95.9 to 98.3) |
| Male | NP | 871 | 13.1 | 75.4 (66.5 to 83.0) | 100 (99.5 to 100) | 100 (95.8 to 100) | 96.4 (94.9 to 97.6) |
| | OP-N | 830 | 11.2 | 79.6 (69.9 to 87.2) | 99.7 (99.0 to 100) | 97.4 (90.8 to 99.7) | 97.5 (96.1 to 98.5) |
| Age [years] [#]: | | | | | | | |
| ≥16 to ≤40 | NP | 1029 | 10.4 | 75.7 (66.5 to 83.5) | 99.9 (99.4 to 100) | 98.8 (93.4 to 100) | 97.3 (96.0 to 98.2) |
| | OP-N | 1037 | 8.9 | 69.8 (59.6 to 78.7) | 99.8 (99.3 to 100) | 97.1 (89.9 to 99.6) | 97.1 (95.9 to 98.1) |
| >40 to ≤65 | NP | 612 | 14.1 | 76.7 (66.4 to 85.2) | 99.6 (98.6 to 100) | 97.1 (89.9 to 99.6) | 96.3 (94.4 to 97.7) |
| | OP-N | 521 | 11.1 | 79.3 (66.6 to 88.8) | 99.8 (98.8 to 100) | 97.9 (88.7 to 99.9) | 97.5 (95.6 to 98.7) |
| >65 | NP | 124 | 17.7 | 59.1 (36.4 to 79.3) | 100 (96.4 to 100) | 100 (75.3 to 100) | 91.9 (85.2 to 96.2) |
| | OP-N | 91 | 9.9 | 100 (66.4 to 100) | 100 (95.6 to 100) | 100 (66.4 to 100) | 100 (95.6 to 100) |
| Testing indication[¦]: | | | | | | | |
| symptomatic | NP | 952 | 13.3 | 88.2 (81.3 to 93.2) | 99.8 (99.1 to 100) | 98.2 (93.8 to 99.8) | 98.2 (97.1 to 99.0) |
| | OP-N | 759 | 12.1 | 81.5 (72.1 to 88.9) | 99.9 (99.2 to 100) | 98.7 (92.9 to 100) | 97.5 (96.0 to 98.5) |
| close contact with SARS-CoV-2 infected individual | NP | 688 | 11.5 | 55.7 (44.1 to 66.9) | 99.8 (99.1 to 100) | 97.8 (88.2 to 99.9) | 94.6 (92.5 to 96.2) |
| | OP-N | 752 | 8.0 | 63.3 (49.9 to 75.4) | 99.9 (99.2 to 100) | 97.4 (86.5 to 99.9) | 96.9 (95.4 to 98.1) |
| **PanBio (Abbott), n (NP) = 970** | | | | | | | |
| Vaccinated (at least 1 vaccination)[@]: | | | | | | | |
| Yes | NP | 167 | 7.8 | 69.2 (38.6 to 90.9) | 100.0 (97.6 to 100.0) | 100.0 (66.4 to 100.0) | 97.5 (93.6 to 99.3) |
| No | NP | 1817 | 8.5 | 68.8 (60.9 to 76.0) | 99.9 (99.7 to 100.0) | 99.1 (94.9 to 100.0) | 97.2 (96.3 to 97.9) |
| Previous SARS-CoV-2 infection[§]: | | | | | | | |
| Yes | NP | 134 | 11.9 | 62.5 (35.4 to 84.8) | 100.0 (96.9 to 100.0) | 100.0 (69.2 to 100.0) | 95.2 (89.8 to 98.2) |
| No | NP | 1850 | 8.1 | 70.0 (62.0 to 77.2) | 99.9 (99.7 to 100.0) | 99.1 (94.9 to 100.0) | 97.4 (96.6 to 98.1) |
| Sex[†]: | | | | | | | |
| Female | NP | 1075 | 7.1 | 69.7 (58.1 to 79.8) | 99.9 (99.4 to 100) | 98.1 (90.1 to 100) | 97.7 (96.6 to 98.6) |

| | | | | | | | |
|---|---|---|---|---|---|---|---|
| Male | NP | 977 | 9.9 | 68.0 (57.8 to 77.1) | 100 (99.6 to 100) | 100 (94.6 to 100) | 96.6 (95.2 to 97.7) |
| Age [years] [#]: | | | | | | | |
| ≥16 to ≤40 | NP | 1307 | 7.5 | 61.2 (50.8 to 70.9) | 100 (99.7 to 100) | 100 (94.0 to 100) | 97.0 (95.8 to 97.8) |
| >40 to ≤65 | NP | 635 | 10.9 | 79.7 (68.3 to 88.4) | 99.8 (99.0 to 100) | 98.2 (90.4 to 100) | 97.6 (96.0 to 98.7) |
| >65 | NP | 112 | 5.4 | 66.7 (22.3 to 95.7) | 100 (96.6 to 100) | 100 (39.8 to 100) | 98.1 (93.5 to 99.8) |
| Testing indication[!]: | | | | | | | |
| symptomatic | NP | 1273 | 7.5 | 70.8 (60.7 to 79.7) | 99.9 (99.5 to 100) | 98.6 (92.2 to 100) | 97.7 (96.7 to 98.4) |
| close contact with SARS-CoV-2 infected individual | NP | 594 | 9.9 | 64.4 (50.9 to 76.4) | 100 (99.3 to 100) | 100 (90.7 to 100) | 96.2 (94.3 to 97.6) |

NP = deep nasopharyngeal; OP-N = combined oropharyngeal and nasal sampling; PPV=positive predictive value; NPV=negative predictive value.

*SARS-CoV-2 infection based on molecular test result.

[@] COVID-19 vaccination status not available from 34, 14, 53, and 72 participants, including 7, 0, 4, and 7 with a positive molecular test result, in the BD-Veritor group, SD-Biosensor NP group, SD-Biosensor OP-N group, and PanBio NP group, respectively.

[§] Previous SARS-CoV-2 infection information not available from 48, 14, 56, and 72 participants in the BD-Veritor group, SD-Biosensor NP group, SD-Biosensor OP-N group, and PanBio NP group, respectively.

[†] Sex not available from 6, 4, 3, and 4 participants in the BD-Veritor group, SD-Biosensor NP group, SD-Biosensor OP-N group, and PanBio NP group, respectively.

[#] Age was not available from 3, 4, 4, and 2 participants in the BD-Veritor group, SD-Biosensor NP group, SD-Biosensor OP-N group, and PanBio NP group, respectively.

[!] Indication for testing was referral for other reason for 73, 92, 93, and 91 participants and unknown for 67, 37, 85, and 98 participants in the BD-Veritor group, SD-Biosensor NP group, SD-Biosensor OP-N group, and PanBio NP group, respectively.

**Table S4** Two-by-two tables used in primary and secondary analysis to determine diagnostic accuracy parameters of the BD Veritor$^{TM}$ System by Becton Dickinson ('BD-Veritor') using routine sampling (OP-N)

| Analysis | Subgroup | | | | |
|---|---|---|---|---|---|
| **Primary** | | | **Ref+** | **Ref-** | **Total** |
| | | **Test+** | **129** | **2** | **131** |
| | | **Test-** | **59** | **1251** | **1310** |
| | | **Total** | **188** | **1253** | **1441** |
| **Secondary (stratified):** | | | | | |
| Infectiousness viral load cut-off¶ | | | Ref+ | Ref- | Total |
| | | Test+ | 125 | 6 | 131 |
| | | Test- | 21 | 1289 | 1310 |
| | | Total | 146 | 1295 | 1441 |
| Symptoms present at sampling‡: | Yes | | Ref+ | Ref- | Total |
| | | Test+ | 91 | 1 | 92 |
| | | Test- | 29 | 541 | 570 |
| | | Total | 120 | 542 | 662 |
| | No | | Ref+ | Ref- | Total |
| | | Test+ | 33 | 1 | 34 |
| | | Test- | 26 | 682 | 708 |
| | | Total | 59 | 683 | 742 |
| Vaccinated (at least 1 vaccination)[@]: | Yes | | Ref+ | Ref- | Total |
| | | Test+ | 10 | 0 | 10 |
| | | Test- | 6 | 136 | 142 |
| | | Total | 16 | 136 | 152 |
| | No | | Ref+ | Ref- | Total |
| | | Test+ | 113 | 2 | 115 |
| | | Test- | 49 | 1091 | 1140 |
| | | Total | 162 | 1093 | 1255 |
| Previous SARS-CoV-2 infection[§]: | Yes | | Ref+ | Ref- | Total |
| | | Test+ | 7 | 0 | 7 |
| | | Test- | 4 | 91 | 95 |
| | | Total | 11 | 91 | 102 |
| | No | | Ref+ | Ref- | Total |
| | | Test+ | 115 | 2 | 117 |
| | | Test- | 49 | 1125 | 1174 |
| | | Total | 164 | 1127 | 1291 |
| Sex[†]: | Female | | Ref+ | Ref- | Total |
| | | Test+ | 68 | 1 | 69 |
| | | Test- | 34 | 695 | 729 |
| | | Total | 102 | 696 | 798 |
| | Male | | Ref+ | Ref- | Total |
| | | Test+ | 61 | 1 | 62 |
| | | Test- | 25 | 550 | 575 |
| | | Total | 86 | 551 | 637 |
| Age [years] [#]: | ≥16 to ≤40 | | Ref+ | Ref- | Total |
| | | Test+ | 61 | 0 | 61 |
| | | Test- | 25 | 661 | 686 |
| | | Total | 86 | 661 | 747 |
| | >40 to ≤65 | | Ref+ | Ref- | Total |
| | | Test+ | 60 | 0 | 60 |
| | | Test- | 29 | 474 | 503 |
| | | Total | 89 | 474 | 563 |
| | >65 | | Ref+ | Ref- | Total |
| | | Test+ | 8 | 2 | 10 |
| | | Test- | 5 | 113 | 118 |
| | | Total | 13 | 115 | 128 |
| Testing indication[!]: | symptomatic | | Ref+ | Ref- | Total |
| | | Test+ | 58 | 2 | 60 |
| | | Test- | 22 | 419 | 441 |
| | | Total | 80 | 421 | 501 |

| | | Ref+ | Ref- | Total |
|---|---|---|---|---|
| close contact with SARS-CoV-2 infected individual | Test+ | 64 | 0 | 64 |
| | Test- | 29 | 707 | 736 |
| | Total | 93 | 707 | 800 |

Ref = molecular reference standard test; Test = BD Veritor$^{TM}$ System by Becton Dickinson

¶Viral load cut-off for infectiousness, defined as viral load above which 95% of people with a positive RT-PCR test result had a positive viral culture[1] was 5.2 log10 SARS-CoV-2 E-gene copies/mL

‡Symptoms not available for 37 participants.

[@] COVID-19 vaccination status not available from 34 participants.

[§] Previous SARS-CoV-2 infection information not available from 48 participants.

[†] Sex not available from 6 participants.

[#] Age was not available from 7 participants.

[!] Indication for testing was referral for other reason for 73 participants and unknown for 67 participants.

**Table S5** Two-by-two tables used in primary and secondary analysis to determine diagnostic accuracy parameters of the Roche/SD Biosensor by Roche Diagnostics (‘SD-Biosensor’) using routine sampling (NP)

| Analysis | Subgroup | | | | |
|---|---|---|---|---|---|
| **Primary** | | | **Ref+** | **Ref-** | **Total** |
| | | **Test+** | **160** | **3** | **163** |
| | | **Test-** | **55** | **1551** | **1606** |
| | | **Total** | **215** | **1554** | **1769** |
| **Secondary (stratified):** | | | | | |
| Infectiousness viral load cut-off¶ | | | Ref+ | Ref- | Total |
| | | Test+ | 160 | 3 | 163 |
| | | Test- | 22 | 1584 | 1606 |
| | | Total | 182 | 1587 | 1769 |
| Symptoms present at sampling‡: | Yes | | Ref+ | Ref- | Total |
| | | Test+ | 126 | 2 | 128 |
| | | Test- | 25 | 938 | 963 |
| | | Total | 151 | 940 | 1091 |
| | No | | Ref+ | Ref- | Total |
| | | Test+ | 34 | 1 | 35 |
| | | Test- | 29 | 594 | 623 |
| | | Total | 63 | 595 | 658 |
| Vaccinated (at least 1 vaccination)[@]: | Yes | | Ref+ | Ref- | Total |
| | | Test+ | 9 | 0 | 9 |
| | | Test- | 1 | 86 | 87 |
| | | Total | 10 | 86 | 96 |
| | No | | Ref+ | Ref- | Total |
| | | Test+ | 151 | 3 | 154 |
| | | Test- | 54 | 1451 | 1505 |
| | | Total | 205 | 1454 | 1659 |
| Previous SARS-CoV-2 infection[§]: | Yes | | Ref+ | Ref- | Total |
| | | Test+ | 13 | 0 | 13 |
| | | Test- | 6 | 168 | 174 |
| | | Total | 19 | 168 | 187 |
| | No | | Ref+ | Ref- | Total |
| | | Test+ | 147 | 3 | 150 |
| | | Test- | 49 | 1369 | 1418 |
| | | Total | 196 | 1372 | 1568 |
| Sex[†]: | Female | | Ref+ | Ref- | Total |
| | | Test+ | 74 | 3 | 77 |
| | | Test- | 27 | 790 | 817 |
| | | Total | 101 | 793 | 894 |
| | Male | | Ref+ | Ref- | Total |
| | | Test+ | 86 | 0 | 86 |
| | | Test- | 28 | 757 | 785 |
| | | Total | 114 | 757 | 871 |
| Age [years] [#]: | ≥16 to ≤40 | | Ref+ | Ref- | Total |
| | | Test+ | 81 | 1 | 82 |
| | | Test- | 26 | 921 | 947 |
| | | Total | 107 | 922 | 1029 |
| | >40 to ≤65 | | Ref+ | Ref- | Total |
| | | Test+ | 66 | 2 | 68 |
| | | Test- | 20 | 524 | 544 |
| | | Total | 86 | 526 | 612 |
| | >65 | | Ref+ | Ref- | Total |
| | | Test+ | 13 | 0 | 13 |
| | | Test- | 9 | 102 | 111 |
| | | Total | 22 | 102 | 124 |
| Testing indication[ǀ]: | symptomatic | | Ref+ | Ref- | Total |
| | | Test+ | 112 | 2 | 114 |
| | | Test- | 15 | 823 | 838 |
| | | Total | 127 | 825 | 952 |

| | | Ref+ | Ref- | Total |
|---|---|---|---|---|
| close contact with SARS-CoV-2 infected individual | Test+ | 44 | 1 | 45 |
| | Test- | 35 | 608 | 643 |
| | Total | 79 | 609 | 688 |

Ref = molecular reference standard test; Test = Roche/SD Biosensor by Roche Diagnostics

¶Viral load cut-off for infectiousness, defined as viral load above which 95% of people with a positive RT-PCR test result had a positive viral culture[1] was 5.2 log10 SARS-CoV-2 E-gene copies/mL.

‡Symptoms not available for 20 participants.

[@] COVID-19 vaccination status not available from 14 participants.

[§] Previous SARS-CoV-2 infection information not available from 14 participants.

[†] Sex not available from 4 participants.

[#] Age was not available from 13 participants.

[!] Indication for testing was referral for other reason for 92 participants and unknown for 37 participants.

**Table S6** Two-by-two tables used in primary and secondary analysis to determine diagnostic accuracy parameters of the Roche/SD Biosensor by Roche Diagnostics ('SD-Biosensor') using less invasive sampling (OP-N)

| Analysis | Subgroup | | | | |
|---|---|---|---|---|---|
| **Primary** | | | **Ref+** | **Ref-** | **Total** |
| | | **Test+** | **123** | **3** | **126** |
| | | **Test-** | **41** | **1522** | **1563** |
| | | **Total** | **164** | **1525** | **1689** |
| **Secondary (stratified):** | | | | | |
| Infectiousness viral load cut-off¶ | | | Ref+ | Ref- | Total |
| | | Test+ | 118 | 8 | 126 |
| | | Test- | 23 | 1540 | 1563 |
| | | Total | 141 | 1548 | 1689 |
| Symptoms present at sampling‡: | Yes | | Ref+ | Ref- | Total |
| | | Test+ | 90 | 2 | 92 |
| | | Test- | 24 | 784 | 808 |
| | | Total | 114 | 786 | 900 |
| | No | | Ref+ | Ref- | Total |
| | | Test+ | 29 | 0 | 29 |
| | | Test- | 17 | 689 | 706 |
| | | Total | 46 | 689 | 735 |
| Vaccinated (at least 1 vaccination)[@]: | Yes | | Ref+ | Ref- | Total |
| | | Test+ | 14 | 0 | 14 |
| | | Test- | 3 | 207 | 210 |
| | | Total | 17 | 207 | 224 |
| | No | | Ref+ | Ref- | Total |
| | | Test+ | 105 | 2 | 107 |
| | | Test- | 38 | 1267 | 1305 |
| | | Total | 143 | 1269 | 1412 |
| Previous SARS-CoV-2 infection[§]: | Yes | | Ref+ | Ref- | Total |
| | | Test+ | 6 | 1 | 7 |
| | | Test- | 5 | 184 | 189 |
| | | Total | 11 | 185 | 196 |
| | No | | Ref+ | Ref- | Total |
| | | Test+ | 113 | 1 | 114 |
| | | Test- | 36 | 1287 | 1323 |
| | | Total | 149 | 1288 | 1437 |
| Sex[†]: | Female | | Ref+ | Ref- | Total |
| | | Test+ | 48 | 1 | 49 |
| | | Test- | 22 | 785 | 807 |
| | | Total | 70 | 786 | 856 |
| | Male | | Ref+ | Ref- | Total |
| | | Test+ | 74 | 2 | 76 |
| | | Test- | 19 | 735 | 754 |
| | | Total | 93 | 737 | 830 |
| Age [years] [#]: | ≥16 to ≤40 | | Ref+ | Ref- | Total |
| | | Test+ | 67 | 2 | 69 |
| | | Test- | 29 | 975 | 1004 |
| | | Total | 96 | 977 | 1073 |
| | >40 to ≤65 | | Ref+ | Ref- | Total |
| | | Test+ | 46 | 1 | 47 |
| | | Test- | 12 | 462 | 474 |
| | | Total | 58 | 463 | 521 |
| | >65 | | Ref+ | Ref- | Total |
| | | Test+ | 9 | 0 | 9 |
| | | Test- | 0 | 82 | 82 |
| | | Total | 9 | 82 | 91 |
| Testing indication[!]: | symptomatic | | Ref+ | Ref- | Total |
| | | Test+ | 75 | 1 | 76 |
| | | Test- | 17 | 666 | 683 |
| | | Total | 92 | 667 | 759 |

| | | Ref+ | Ref- | Total |
|---|---|---|---|---|
| close contact with SARS-CoV-2 infected individual | Test+ | 38 | 1 | 39 |
| | Test- | 22 | 691 | 713 |
| | Total | 60 | 692 | 752 |

Ref = molecular reference standard test; Test = Roche/SD Biosensor by Roche Diagnostics
¶Viral load cut-off for infectiousness, defined as viral load above which 95% of people with a positive RT-PCR test result had a positive viral culture[1] was 5.2 log10 SARS-CoV-2 E-gene copies/mL.
‡Symptoms not available for 53 participants.
[@] COVID-19 vaccination status not available from 53 participants.
[§] Previous SARS-CoV-2 infection information not available from 56 participants.
[†] Sex not available from 3 participants.
[#] Age was not available from 13 participants.
[!] Indication for testing was referral for other reason for 93 participants and unknown for 85 participants.

**Table S7** Two-by-two tables used in primary and secondary analysis to determine diagnostic accuracy parameters of the PanBio by Abbot ('PanBio') using routine sampling (NP)

| Analysis | Subgroup | | | | |
|---|---|---|---|---|---|
| **Primary** | | | **Ref+** | **Ref-** | **Total** |
| | | **Test+** | **119** | **1** | **120** |
| | | **Test-** | **54** | **1882** | **1936** |
| | | **Total** | **173** | **1883** | **2056** |
| **Secondary (stratified):** | | | | | |
| Infectiousness viral load cut-off¶[$] | | | Ref+ | Ref- | Total |
| | | Test+ | 108 | 2 | 110 |
| | | Test- | 13 | 1916 | 1929 |
| | | Total | 121 | 1918 | 2039 |
| Symptoms present at sampling‡: | Yes | | Ref+ | Ref- | Total |
| | | Test+ | 96 | 1 | 97 |
| | | Test- | 37 | 1336 | 1373 |
| | | Total | 133 | 1337 | 1470 |
| | No | | Ref+ | Ref- | Total |
| | | Test+ | 19 | 0 | 19 |
| | | Test- | 15 | 477 | 492 |
| | | Total | 34 | 477 | 511 |
| Vaccinated (at least 1 vaccination)[@]: | Yes | | Ref+ | Ref- | Total |
| | | Test+ | 9 | 0 | 9 |
| | | Test- | 4 | 154 | 158 |
| | | Total | 13 | 154 | 167 |
| | No | | Ref+ | Ref- | Total |
| | | Test+ | 106 | 1 | 107 |
| | | Test- | 48 | 1662 | 1710 |
| | | Total | 154 | 1663 | 1817 |
| Previous SARS-CoV-2 infection[§]: | Yes | | Ref+ | Ref- | Total |
| | | Test+ | 10 | 0 | 10 |
| | | Test- | 6 | 118 | 124 |
| | | Total | 16 | 118 | 134 |
| | No | | Ref+ | Ref- | Total |
| | | Test+ | 105 | 1 | 106 |
| | | Test- | 45 | 1699 | 1744 |
| | | Total | 150 | 1700 | 1850 |
| Sex[†]: | Female | | Ref+ | Ref- | Total |
| | | Test+ | 53 | 1 | 54 |
| | | Test- | 23 | 998 | 1021 |
| | | Total | 76 | 999 | 1075 |
| | Male | | Ref+ | Ref- | Total |
| | | Test+ | 66 | 0 | 66 |
| | | Test- | 31 | 880 | 911 |
| | | Total | 97 | 880 | 977 |
| Age [years] [#]: | ≥16 to ≤40 | | Ref+ | Ref- | Total |
| | | Test+ | 60 | 0 | 60 |
| | | Test- | 38 | 1209 | 1247 |
| | | Total | 98 | 1209 | 1307 |
| | >40 to ≤65 | | Ref+ | Ref- | Total |
| | | Test+ | 55 | 1 | 56 |
| | | Test- | 14 | 565 | 579 |
| | | Total | 69 | 566 | 635 |
| | >65 | | Ref+ | Ref- | Total |
| | | Test+ | 4 | 0 | 4 |
| | | Test- | 2 | 106 | 108 |
| | | Total | 6 | 106 | 112 |
| Testing indication[!]: | symptomatic | | Ref+ | Ref- | Total |
| | | Test+ | 68 | 1 | 69 |
| | | Test- | 28 | 1176 | 1204 |
| | | Total | 96 | 1177 | 1273 |

| | | Ref+ | Ref- | Total |
|---|---|---|---|---|
| close contact with SARS-CoV-2 infected individual | Test+ | 38 | 0 | 38 |
| | Test- | 21 | 535 | 556 |
| | Total | 59 | 535 | 594 |

Ref = molecular reference standard test; Test = PanBio by Abbot

¶Viral load cut-off for infectiousness, defined as viral load above which 95% of people with a positive RT-PCR test result had a positive viral culture[1] was 5.2 log10 SARS-CoV-2 E-gene copies/mL.

[$]Viral load unavailable for 17 participants.

‡Symptoms not available for 75 participants.

[@] COVID-19 vaccination status not available from 72 participants.

[§] Previous SARS-CoV-2 infection information not available from 72 participants.

[†] Sex not available from 4 participants.

[#] Age was not available from 17 participants.

[!] Indication for testing was referral for other reason for 91 participants and unknown for 98 participants.

**Table S8** Two-by-two tables used in primary and secondary analysis to determine diagnostic accuracy parameters of the PanBio by Abbot ('PanBio') using less invasive sampling (OP-N)

| Analysis | Subgroup | | | | |
|---|---|---|---|---|---|
| **Primary** | | | **Ref+** | **Ref-** | **Total** |
| | | **Test+** | **18** | **0** | **18** |
| | | **Test-** | **15** | **937** | **952** |
| | | **Total** | **33** | **937** | **970** |
| **Secondary (stratified):** | | | | | |
| Infectiousness viral load cut-off¶ | | | Ref+ | Ref- | Total |
| | | Test+ | 18 | 0 | 18 |
| | | Test- | 9 | 943 | 952 |
| | | Total | 27 | 943 | 970 |
| Symptoms present at sampling‡: | Yes | | Ref+ | Ref- | Total |
| | | Test+ | 13 | 0 | 13 |
| | | Test- | 7 | 644 | 651 |
| | | Total | 20 | 644 | 664 |
| | No | | Ref+ | Ref- | Total |
| | | Test+ | 3 | 0 | 3 |
| | | Test- | 8 | 277 | 285 |
| | | Total | 11 | 277 | 288 |
| Vaccinated (at least 1 vaccination)[@]: | Yes | | Ref+ | Ref- | Total |
| | | Test+ | 3 | 0 | 3 |
| | | Test- | 3 | 295 | 298 |
| | | Total | 6 | 295 | 301 |
| | No | | Ref+ | Ref- | Total |
| | | Test+ | 13 | 0 | 13 |
| | | Test- | 12 | 627 | 639 |
| | | Total | 25 | 627 | 652 |
| Previous SARS-CoV-2 infection[§]: | Yes | | Ref+ | Ref- | Total |
| | | Test+ | 0 | 0 | 0 |
| | | Test- | 4 | 53 | 57 |
| | | Total | 4 | 53 | 57 |
| | No | | Ref+ | Ref- | Total |
| | | Test+ | 15 | 0 | 15 |
| | | Test- | 11 | 867 | 878 |
| | | Total | 26 | 867 | 893 |
| Sex[†]: | Female | | Ref+ | Ref- | Total |
| | | Test+ | 6 | 0 | 6 |
| | | Test- | 9 | 458 | 467 |
| | | Total | 15 | 458 | 473 |
| | Male | | Ref+ | Ref- | Total |
| | | Test+ | 12 | 0 | 12 |
| | | Test- | 6 | 473 | 479 |
| | | Total | 18 | 473 | 491 |
| Age [years] [#]: | ≥16 to ≤40 | | Ref+ | Ref- | Total |
| | | Test+ | 12 | 0 | 12 |
| | | Test- | 13 | 568 | 581 |
| | | Total | 25 | 568 | 593 |
| | >40 to ≤65 | | Ref+ | Ref- | Total |
| | | Test+ | 6 | 0 | 6 |
| | | Test- | 2 | 309 | 311 |
| | | Total | 8 | 309 | 317 |
| | >65 | | Ref+ | Ref- | Total |
| | | Test+ | 0 | 0 | 0 |
| | | Test- | 0 | 54 | 54 |
| | | Total | 0 | 0 | 0 |
| Testing indication[']: | symptomatic | | Ref+ | Ref- | Total |

| | | | | |
|---|---|---|---|---|
| | Test+ | 12 | 0 | 12 |
| | Test- | 4 | 597 | 601 |
| | Total | 16 | 597 | 6013 |
| close contact with SARS-CoV-2 infected individual | | Ref+ | Ref- | Total |
| | Test+ | 3 | 0 | 3 |
| | Test- | 9 | 224 | 233 |
| | Total | 12 | 224 | 236 |

Ref = molecular reference standard test; Test = PanBio by Abbot

¶Viral load cut-off for infectiousness, defined as viral load above which 95% of people with a positive RT-PCR test result had a positive viral culture[1] was 5.2 log10 SARS-CoV-2 E-gene copies/mL.

‡Symptoms not available for 18 participants.

[@] COVID-19 vaccination status not available from 17 participants.

[§] Previous SARS-CoV-2 infection information not available from 20 participants.

[†] Sex not available from 6 participants.

[#] Age was not available from 10 participants.

[!] Indication for testing was referral for other reason for 90 participants and unknown for 31 participants.

**Table S9** GISAID data contributors acknowledgement table

We gratefully acknowledge the following Authors from the Originating laboratories responsible for obtaining the specimens and the Submitting laboratories where genetic sequence data were generated and shared via the GISAID Initiative, on which this research is based.

| AccessionID | Originating laboratory | Submitting laboratory | Authors |
|---|---|---|---|
| EPI_ISL_5855699 | Dutch COVID-19 response team | Erasmus Medical Center | Bas Oude Munnink, Reina Sikkema, David Nieuwenhuijse, Irina Chestakova, Anne van der Linden, Marjan Boter, Emmanuelle Munger, Corine GeurtsvanKessel, Annemiek van der Eijk, Richard Molenkamp, Marion Koopmans, on behalf of the Dutch national COVID-19 response team. |
| EPI_ISL_2731321, EPI_ISL_2731323, EPI_ISL_2731327, EPI_ISL_2731348 EPI_ISL_2925600-05, EPI_ISL_2955624-26, EPI_ISL_5051430-33, EPI_ISL_5825426-27, EPI_ISL_5846285-86, EPI_ISL_6182027. | Microvida | Microvida | Suzan D. Pas, Jaco J. Verweij, Joep J. J. M. Stohr. |

All submitters of data may be contacted directly via https://www.gisaid.org

**Supplementary Material 1** Short questionnaire (translated from Dutch)

1. What is the reason for testing?
   *Multiple answers possible*
   - ☐ I have (had) Covid-19 like symptoms
   - ☐ I am a close contact of a SARS-CoV-2 infected person
   - Date of last contact: ___ - ___ (day – month)

   I was notified because of:
   - The infected person is a household member □ yes / □ no
   - Received notification by CoronaMelder app (English: Corona notification app) □ yes / □ no
   - Received notification by public health service (by phone or letter) □ yes / □ no
   - Received notification by SARS-CoV-2 infected person □ yes / □ no
   - Other, ………

   - ☐ GP recommended a SARS-CoV-2 test
   - ☐ Travelled to a high-pandemic country
   - ☐ None of the above, but…….

2. Did you receive a Covid-19 vaccination
   - ☐ No
   - ☐ Yes

If yes, which vaccine did you receive? □ Pfizer □ Moderna □ AstraZeneca □ Janssen □ Unknown
Which number of vaccinations did you receive? □ One □ Two

3. Did you have had a positive SARS-CoV-2 test previously? If yes, how long ago?

□ No □ < 2 months □ 2-6 months □ 6-12 months □ >12 months

4. At this moment, do you have any COVID-19 like symptoms?
   - ☐ No ***END OF QUESTIONNAIRE***
   - ☐ Yes

5. What COVID-19 like symptoms do you currently have?
   *Multiple answers possible*
   - ☐ Common cold
   - ☐ Shortness of breath
   - ☐ Fever
   - ☐ Coughing
   - ☐ Loss of taste or smell
   - ☐ Muscle ache
   - ☐ I have other symptoms

6. What was the moment you first experienced these symptoms?
   - ☐ Today
   - ☐ Yesterday
   - ☐ Two days ago
   - ☐ Three or more days ago

**Supplementary Material 2** Specimen collection, SARS-CoV-2 diagnostic testing, and SARS-CoV-2 virus culture procedures

**Specimen collection and SARS-CoV-2 diagnostic testing procedures**

*West Brabant region, Microvida laboratories at Amphia hospital in Breda and Roosendaal: BD-Veritor*

At this location, the BD Veritor$^{TM}$ by Becton Dickinson Ag-RDT ('BD-Veritor') was used for routine SARS-CoV-2 testing extended with additional RT-PCR testing as reference standard test. Two swabs were taken per person at the Public Health Service test site in the Amphia hospital in Breda. For the RT-PCR, a superficial combined oropharyngeal and nasal (OP-N) swab (about 2.5 cm deep from the edge of the nostril) was taken, placed in universal transport media (HiViralTM) with MagnaPure LC lysis- and binding buffer (Roche Diagnostics, The Netherlands), and transported to the Microvida laboratory in Roosendaal. RT-PCR was performed using the cobas ® SARS-CoV-2 test on the cobas ® 8800 platform (Roche Diagnostics International, Rotkreuz, Switzerland). Ct values for the SARS-CoV-2 E-gene were converted to viral load (genome copies/ml) based on an in-house established standard curve. The assay was used in accordance with the manufacturer's instructions; amplification curves and cycle threshold values were interpreted using the manufacturer's interpretation algorithms, which complied with the European in-vitro diagnostic devices directive. For Ag-RDT testing, the second combined OP-N swab was stored in a sterile dry tube immediately after collection and taken to the on-site Microvida laboratory in the Amphia hospital in Breda. A trained laboratory technician performed the Ag-RDT in accordance with the manufacturer's operating procedure within one hour after the specimen was obtained. The system is intended to be used with a digital reader although validated for visual reading.[2] In this study, visual readout was performed independently by two persons. In case of discrepancies, the result from the digital reader was used.

*Rotterdam city region, laboratory Erasmus MC: SD-Biosensor*

At this location, the Roche/SD Biosensor by Roche Diagnostics ('SD-Biosensor') Ag-RDT was used for routine SARS-CoV-2 testing extended with additional RT-PCR testing as the reference standard test. Two swabs were taken per person at the two Public Health service test sites in Rotterdam-Rijnmond region (Rotterdam Airport and Ahoy). For RT-PCR, a deep combined OP-N swab (>5 cm deep from the edge of the nostril) was taken, placed directly in universal transport media (HiViralTM) and transported to the Erasmus MC Viroscience diagnostic laboratory. Routine RT-PCR testing was performed in virus transport medium using the cobas ® SARS-CoV-2 test on the cobas 6800® platform (Roche Diagnostics International, Rotkreuz, Switzerland). The assay was used in accordance with the manufacturer's instructions; amplification curves and cycle threshold values were interpreted using the manufacturer's interpretation algorithms, which complied with the European in-vitro diagnostic devices directive. The second swab was used for on-site Ag-RDT testing, and procedures differed slightly for the two different sampling methods that were evaluated.

<u>SD-Biosensor Ag-RDT evaluation using routine sampling</u>

A nasopharyngeal (NP) swab (>5 cm deep from the edge of the nostril) was taken using the swab included in the SD-Biosensor Ag-RDT kit (REF No. 9901-NCOV-01G; LOT No QCO3020079/Sub:A-2). The test was carried out immediately on-site at the two Public Health Service test sites in Rotterdam following manufacturer's instructions. Interpretation and recording of test results was performed independently by two persons according to the manufacturer's instructions.

<u>SD-Biosensor Ag-RDT evaluation using less invasive sampling</u>

A superficial combined OP-N swab (about 2.5 cm from the edge of the nostril) was taken using the swab included in the SD-Biosensor Ag-RDT kit (REF No. 9901-NCOV-01G; LOT No QCO3020079/Sub:A-2). The test was carried out immediately on-site at the two Public Health Service test sites in Rotterdam following manufacturer's instructions. Interpretation and recording of test results was performed independently by two persons according to the manufacturer's instructions.

*Zwolle region, Laboratory of Medical Microbiology and Infectious Diseases, Isala Hospital: PanBio*

The standard method for SARS-CoV-2 testing in Isala Hospital is by transcription mediated (TMA) amplification on the Hologic Panther system (AptimaTM SARS-CoV2 assay). It was carried out as usual, in parallel with the PanBio Ag-RDT. TMA positive samples were further analysed by RT-PCR to obtain Ct values for viral load calculation. These samples were stored at 4-8 degrees Celsius until processed for RT-PCR with a maximum of 24 hours during the week and 72 hours in the weekend. In case of discrepant results (TMA-positive, RT-PCR negative), the RT-PCR was repeated once due to possible sampling error. If there was still discrepancy between results, a Ct value of 40 was assigned for subsequent analysis (8/173 (4.6%) and 1/33 (3.0%) participants with a positive reference standard test in the routine sampling and less invasive sampling periods, respectively).

Two swabs were taken per person at the Public Health service test site in Zwolle. For TMA/RT-PCR, a deep combined OP-N swab (>5 cm deep from the edge of the nostril) was taken, placed in 1x Tris-EDTA buffer (Sigma-

Aldrich), and transported to the Isala Hospital Laboratory for Medical Microbiology and Infectious Diseases for analysis in Zwolle. The in-house RT-PCR was performed based on Corman et al.[3] RNA extraction was performed on the KingFisher Flex System (ThermoFisher) using the MagMAX Viral/Pathogen Nucleic Acid Isolation Kit with the manufacturer protocol MVP_2Wash_200_Flex_SARS-COV2 (ThermoFisher). RT-PCR and detection was performed with the TaqMan Fast Virus 1-Step Mastermix (ThermoFisher) on the ABI7500 platform (Applied Biosystems). This RT-PCR has one target: the SARS-CoV-2 E-gene. The second swab was used for on-site Ag-RDT testing, and procedures differed slightly for the two different sampling methods that were evaluated.

PanBio Ag-RDT evaluation using routine sampling
A deep NP swab (>5 cm deep from the edge of the nostril) was taken and tested by PanBio Ag-RDT at thePublic Health Service test site in Zwolle. The collected swab was transferred into supplied extraction tubes. A trained laboratory technician performed the Ag-RDT in accordance with the manufacturer's operating procedure within 2 hours after the specimen was obtained. Readout was performed independently by two persons.

PanBio Ag-RDT evaluation using less invasive sampling
A superficial combined OP-N swab (about 2.5 cm from the edge of the nostril) was tested by the PanBio Ag-RDT at the Public Health Service test site in Zwolle. The collected swab was transferred into supplied extraction tubes. A trained laboratory technician performed the Ag-RDT in accordance with the manufacturer's operating procedure within 2 hours after the specimen was obtained. Readout was performed independently by two persons.'

**Viral load calculation**
We used a viral load cut-off as a proxy of infectiousness of ≥5.2 log10 SARS-CoV-2 E-gene copies/mL, which was the viral load cut-off above which 95% of people with a positive molecular test had a positive virus culture in a recent previous study by our group.[1] For that study, the Erasmus MC viroscience laboratory created a standard curve by testing dilutions of a specific quantified E-gene transcript (primary standard) available from the European Virus Archive (EVAg[4]) with the RT-PCR described by Corman et al.[3] The relation between Ct value and number of E-gene copies/ml was determined by linear regression analysis. Based on this calibration curve a secondary standard derived from cell-cultured virus was prepared and quantified. Serial dilutions of this secondary standard were tested by RT-PCR to prepare a secondary standard curve by linear regression.

In the previous study, we determined whether the cobas PCR platform at Microvida laboratory provided comparable data to the cobas PCR platform at Erasmus MC laboratory by having both laboratories test the same SARS-CoV-2 viral load panel obtained from the National Public Health Institute (RIVM). The Ct values generated in the two laboratories corresponded well. In the current study, a third central laboratory (Isala Hospital in Zwolle) participated. This hospital laboratory used the same medium and reagent volumes as the Erasmus MC laboratory. The Erasmus MC laboratory provided a new serial dilution of a virus culture-derived standard provided to the Isala Hospital laboratory, which they tested by RT-PCR in triplicate. Ct values generated by the Isala Hospital laboratory matched the expected Ct values for that standard based on the Ct value obtained by the Erasmus MC laboratory.

Given the above, we concluded that the same formula to convert Ct values into viral loads (copies/ml) could be used for the Zwolle and Rotterdam laboratories This formula was $62.5 * e^{\frac{43.1-Ct}{1.607}}$. The Microvida laboratory used lower medium/reagent volumes per swab (1.8 ml compared to 6 ml in the other two laboratories; a factor $3\frac{1}{3}$) and that conversion formula therefore was $62.5 * e^{\frac{43.1-Ct}{1.607}} / 3\frac{1}{3}$.

**Whole genome sequencing (WGS)**
In discordant cases (Ag-RDT negative and RT-PCR positive cases) with viral loads above the infectiousness cut-off (≥5.2 log10 SARS-CoV-2 E gene copies/mL) whole genome sequencing (WGS) of the primary clinical specimen was performed by Microvida laboratory to determine the SARS-CoV-2 variant. In short, total nucleic acids were extracted using the QIAsymphony DSP virus pathogen midi kit and pathogen complex 400 protocol of the QIAsymphony Sample Processing system (Qiagen, Germany), with an input volume of 400 µL and output volume of 110 µL.Confirmation SARS-CoV-2 viral load was performed[5] with only samples with Ct value <32 selected for further WGS. cDNA was synthesized using LunaScript® RT SuperMix Kit (New Engeland Biolabs, USA) and library preparation was performed using EasySeq™ RC-PCR SARS-CoV-2 Whole Genome Sequencing kit (Nimagen, The Netherlands) according to manufacturer's instructions. Subsequent next generation sequencing (NGS) of 2x150cycles paired end reads was performed on a Miseq (Illumina, The Netherlands) using MiSeq Reagent Micro Kit v2 according to manufacturer's instructions. Data analyses was performed with an in-house workflow using CLCbio Genomic Workbench v21 (Qiagen, Germany), including (a.o.) read-trimming, NC_045512.2 (NCBI genbank) reference based assembly, local re-alignment and variant detection algorithms.

The consensus genome was extracted and positions with a coverage less than 10 reads were replaced with N. The sequences were manually curated. Genomes with >70% genome coverage were included for lineage assignment using Pangolin (https://pangolin.cog-uk.io/).[6] All genomes with 100% N-gen coverage were used for N-gen mutation analysis using CLCBio Genomic workbench v21 and Nextclade Web 1.7.1 (https://clades.nextstrain.org/).[7] Genomes with >90% genome coverage were uploaded to GISAID (https://www.gisaid.org/).[8] Mutations in the N-gene with a prevalence within their lineage of <10% (according to the mutation tracker tool of https://www.outbreak.info) were reported.[9]
